# SR-TWAS: Leveraging Multiple Reference Panels to Improve TWAS Power by Ensemble Machine Learning

**DOI:** 10.1101/2023.06.20.23291605

**Authors:** Randy L. Parrish, Aron S. Buchman, Shinya Tasaki, Yanling Wang, Denis Avey, Jishu Xu, Philip L. De Jager, David A. Bennett, Michael P. Epstein, Jingjing Yang

## Abstract

Multiple reference panels of a given tissue or multiple tissues often exist, and multiple regression methods could be used for training gene expression imputation models for TWAS. To leverage expression imputation models (i.e., base models) trained with multiple reference panels, regression methods, and tissues, we develop a Stacked Regression based TWAS (SR-TWAS) tool which can obtain optimal linear combinations of base models for a given validation transcriptomic dataset. Both simulation and real studies showed that SR-TWAS improved power, due to increased effective training sample sizes and borrowed strength across multiple regression methods and tissues. Leveraging base models across multiple reference panels, tissues, and regression methods, our real application studies identified 6 independent significant risk genes for Alzheimer’s disease (AD) dementia for supplementary motor area tissue and 9 independent significant risk genes for Parkinson’s disease (PD) for substantia nigra tissue. Relevant biological interpretations were found for these significant risk genes.

## Introduction

Two-stage transcriptome-wide association studies (TWAS) have been widely used in genetic studies of complex traits due to the convenience of using publicly-available transcriptomic reference panels and summary-level genome-wide association study (GWAS) datasets^1–5^. The standard two-stage TWAS method^6,7^ first trains gene expression imputation models (per gene per tissue) using a transcriptomic reference panel in Stage I, taking quantitative gene expression traits as response variables and nearby (cis-) or genome-wide (cis- and trans-) genetic variants as predictors. The non-zero genetic effect sizes estimated in the gene expression imputation models are considered effect sizes of a broad sense of expression quantitative trait loci (eQTL), which are taken as variant weights to conduct gene-based association tests with GWAS data (individual-level or summary-level) in Stage II.

Various TWAS techniques have been developed, employing diverse regression methods to train models for imputing gene expression. Additionally, multiple transcriptomic reference panels are made available to the public and could be used in TWAS. Consequently, it is possible to train multiple gene expression imputation models by employing distinct regression methods, employing multiple transcriptomic reference panels of the same tissue type, or utilizing transcriptomic data from multiple tissues within a given reference panel. For example, multiple regression methods, such as penalized regression with Elastic-Net penalty (used by PrediXcan^7,8^) and nonparametric Bayesian Dirichlet process regression (DPR) model (used by TIGAR^9^), have trained gene expression imputation models using the same Genotype-Tissue Expression (GTEx)^10^ V8 reference data of 48 human tissue types. The Religious Orders Study (ROS)^11^, Rush Memory and Aging Project (MAP)^11^, and the GTEx^10^ V8 project all profile transcriptomic data of prefrontal cortex (PFC) brain tissue and genome-wide genetic data of the same samples, providing multiple reference panels of PFC tissue for TWAS. Thus, leveraging multiple trained gene expression imputation models of the same target gene across multiple regression methods, multiple reference panels, and multiple tissue types is expected to improve TWAS power, for more robustly modeling the unknown genetic architecture of the target gene expression by multiple regression models, having an increased training sample size with multiple reference panels, or borrowing strength across multiple tissue types with correlated gene expression.

Multiple approaches that can take advantage of transcriptomic reference data for multiple tissues and/or multiple reference panels have been developed. For example, UTMOST uses group LASSO-penalized multivariate regression to impute cross-tissue expression^12^. TisCoMM uses the same multivariate regression model for gene expression prediction models for leveraging gene expression across multiple tissues, but utilizes a unified probabilistic model to test the overall and tissue-specific gene-trait associations^13^. SWAM estimates a vector of weights for input expression imputation models such that the weighted average of the input models will give the lowest mean squared error with respect to individual-level reference expression of the target tissue^14^. However, these approaches have drawbacks such as requiring individual-level reference data, being computationally expensive, and user-unfriendly. For example, UTMOST and TisCoMM require individual-level reference data for all tissues to train gene expression imputation models. In order to control for multicollinearity, SWAM considers a regularization parameter which requires fine-tuning based on the covariance structure of Genetically Regulated gene eXpression (GReX) of all considered tissues which must be derived using individual-level reference transcriptomic data^14^. Additionally, SWAM requires that the input of trained gene expression imputation models in the same SQL database format as used for PrediXcan output^14^.

To fill this gap, we develop a novel TWAS method to leverage multiple summary-level gene expression imputation models (i.e., base models) trained for the same target gene by the ensemble machine learning technique of stacked regression^15,16^. We refer to this novel TWAS method as Stacked Regression-based TWAS (i.e., SR-TWAS). SR-TWAS first uses a validation transcriptomic dataset of the target tissue type to optimally train a set of weights for the multiple expression imputation base models per target gene (Stage I), by optimizing the gene expression prediction *R*^2^ (i.e., the squared correlation between observed and predicted gene expression levels) in the validation dataset. Then SR-TWAS takes the weighted average eQTL effect sizes as the corresponding variant weights for gene-based association tests in Stage II. The trained expression imputation models by SR-TWAS are specific for the tissue type of the validation data, and the identified TWAS risk genes are interpreted with potential genetic effects mediated through their gene expression of the validation tissue type.

In the following sections, we first briefly describe the stacked regression method used by SR-TWAS and the GTEx V8 and ROS/MAP reference transcriptomic datasets used in this study. Then we describe the results of our simulation studies, validation studies using the real reference data, as well as application TWAS of AD dementia and PD. Last, we end with a discussion.

## Results

### Overview of SR-TWAS

In the framework of TWAS^7,9,17^, a multiple linear regression model is assumed for training gene expression imputation models, taking quantitative gene expression levels *E_g_* of the target gene and tissue as the response variable and cis-acting genetic variants nearby the target gene region (genotype matrix *G*) as predictors, as shown in the following formula:

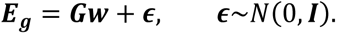

The eQTL effect sizes ***w*** could be trained by different regression methods and/or using different reference panels with matched expression and genotype data (*E_g_*, *G*).

Assume there are a total of *K* base gene expression imputation models trained for the same target gene, with 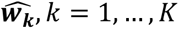. Let **E***_vg_* denote the gene expression levels of the target gene *g* for the target tissue type in the validation data, and **G***_v_* denote the genotype matrix of genetic predictors in the validation data. Then the predicted GReX of the validation samples by the *k*th base model are given by ***G**_v_**ŵ**_k_*. The stacked regression method^15,16^ will solve for a set of optimal base model weights *ζ_1_*,…, *ζ_K_*, by maximizing the regression *R*^2^ between the profiled gene *k=1* expression *E_vg_* and the weighted average GReX, 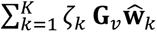, of *K* base models, i.e., minimizing the following loss function of 1 *− R*^2^:

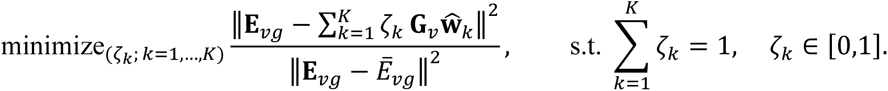

As a result, we will obtain a set of model weights *ζ_k_* for *k* = 1, …, *K* base models, and a set of eQTL effect sizes *w∼* given by the weighted average of the eQTL effect sizes of *K* base models, 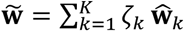 (Stage I). Then the final predicted GReX for test genotype data **G*_t_*** is given by 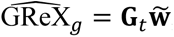, and *w∼* will be taken as variant weights in the gene-based association tests by SR-TWAS in Stage II. Genes with 5-fold cross validation (CV) *R*^2^ > 0.5% in the validation dataset by SR-TWAS are considered as having a valid imputation model and will be tested in Stage II^9^. Here, *w∼* is the trained eQTL effect sizes by SR-TWAS (Stage I) for the target gene of the tissue of the validation data and identified significant genes from Stage II have potential genetic effects mediated through the transcriptome of the tissue of the validation data.

### Reference Transcriptomic Data

#### GTEx V8

The Genotype-Tissue Expression project version 8 (GTEx V8) release provides a comprehensive reference dataset of Whole Genome Sequencing (WGS) genotype data with matched RNA-seq transcriptomic data from 54 non-diseased tissues of 838 post-mortem donors of European ancestry^10^. For our real data analysis, we used publicly-available GReX imputation models trained by TIGAR-V2^9^ and PrediXcan^7,8,18^ with GTEx V8 reference data for European subjects as base models. Supplementary Table 1 shows the disease status of GTEx subjects for tissues used in the AD or PD TWAS. Subjects in the GTEx V8 cohort are generally healthy. AD or other dementia was reported for only 4% of subjects, with missing data for ∼1% subjects. The majority of GTEx subjects used in PD TWAS were reported to be without PD, with <1% reported with PD and with missing data for 13% of training and 6% of validation subjects.

#### ROS/MAP

The Religious Orders Study (ROS) and Rush Memory and Aging Project (MAP) are two longitudinal prospective clinical-pathologic cohort studies of aging and AD, which are collectively referred to as ROS/MAP^11^. ROS recruits participants from religious orders across the U.S. while MAP recruits lay persons in the northeastern Illinois area^11^. All participants in both studies are recruited without known dementia and agree to annual clinical evaluations and brain donation upon death^11^. Only subjects of European ancestry were used in this study. Disease status of ROS/MAP participants used in this study are shown in Supplementary Table 1. Of the ROS/MAP subjects used in the training data of AD TWAS, 57% had normal cognitive function, 40% had AD or other dementia, and 4% had missing disease status. Of the ROS/MAP subjects used in the validation data of AD TWAS, 75% had normal cognitive function and 25% had AD or other types of dementia.

### Simulation Studies

#### Simulation study design

We used the real genotype data of gene *ABCA7* from ROS/MAP and GTEx V8 to simulate gene expression and phenotypes, and considered multiple scenarios with varying proportions of causal SNPs (*p_causal_* = (0.001, 0.01, 0.05, 0.1)) and gene expression heritability (i.e., the proportion of gene expression variation due to genetics, *h_e_*^2^ = (0.1, 0.2, 0.5)). We randomly selected n=465 training samples with Whole Genome Sequencing (WGS) genotype data from the ROS/MAP cohort and GTEx V8 cohort, respectively. We randomly selected n=400 and n=800 samples with WGS genotype data from ROS/MAP as our validation and test cohorts, respectively. ROS/MAP training, validation, and test samples were simulated with the same causal SNPs (i.e., eQTL), while training samples from the GTEx V8 cohort were simulated with true causal SNPs that were 50% overlapped with the ones for ROS/MAP samples. The simulated expression heritability was the same for both ROS/MAP and GTEx V8 samples.

We compared the performance of SR-TWAS with a Naïve approach which takes the average of base models as the trained gene expression imputation model, that is, taking 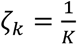 *k* = 1, …, *K*. Two base models per gene were respectively trained by PrediXcan (penalized regression with Elastic-Net penalty; PrediXcan-GTEx) with the GTEx training samples (n=465), and by TIGAR (nonparametric Bayesian Dirichlet process regression; TIGAR-ROSMAP) with the ROS/MAP training samples (n=465). SR-TWAS and Naïve models were then obtained by using these trained base models. Validation data (*n* = 400) were used to train SR-TWAS models and an additional TIGAR base model (TIGAR_ROSMAP_valid). The SR-TWAS and TIGAR_ROSMAP_valid models were then averaged to create a new model (Avg-valid+SR). Gene expression imputation models (by SR-TWAS and Naïve methods) with 5-fold cross-validation *R*^2^ > 0.5% in the validation cohort were considered valid models, which were used to produce the Avg-valid-SR models and used in the follow-up gene-based association tests. Test data (n=800) were used for assessing GReX prediction performance and TWAS power, with 1,000 repeated simulations per scenario. We compared the performance of three base models of the training and validation data (PrediXcan-GTEx, TIGAR-ROSMAP, and TIGAR-ROSMAP_valid), Naïve method, SR-TWAS, and Avg-valid+SR models.

#### Simulation study results

As shown in **Fig. 1**, we showed that Avg-valid+SR obtained the highest test *R*^2^ for gene expression imputation across 11 of 12 scenarios, and the SR-TWAS models had the second-best performance. SR-TWAS obtained the highest test *R*^2^ for gene expression imputation in the *p*_causal_ = 0.01 and *h*^2^ = 0.5 scenario, where it slightly outperformed the averaged models. The base models trained by TIGAR with ROS/MAP training and validation samples (TIGAR-ROSMAP, TIGAR-ROSMAP_valid) performed similarly well, because the test data were generated under the same model assumptions as the ROS/MAP training and validation data. Here, the Avg-valid+SR models performed best for leveraging the predictive information provided by all three base models. The Naïve method and PrediXcan-GTEx base models did not perform well because the PrediXcan-GTEx base models were trained using GTEx training data which only shared half of the true causal SNPs as the validation and test data. The Naïve approach of taking averages of the PrediXcan-GTEx and TIGAR-ROSMAP base models had poor performance because of the heterogeneous genetic architecture between the GTEx training cohort and test cohort.

**Fig. 1.**
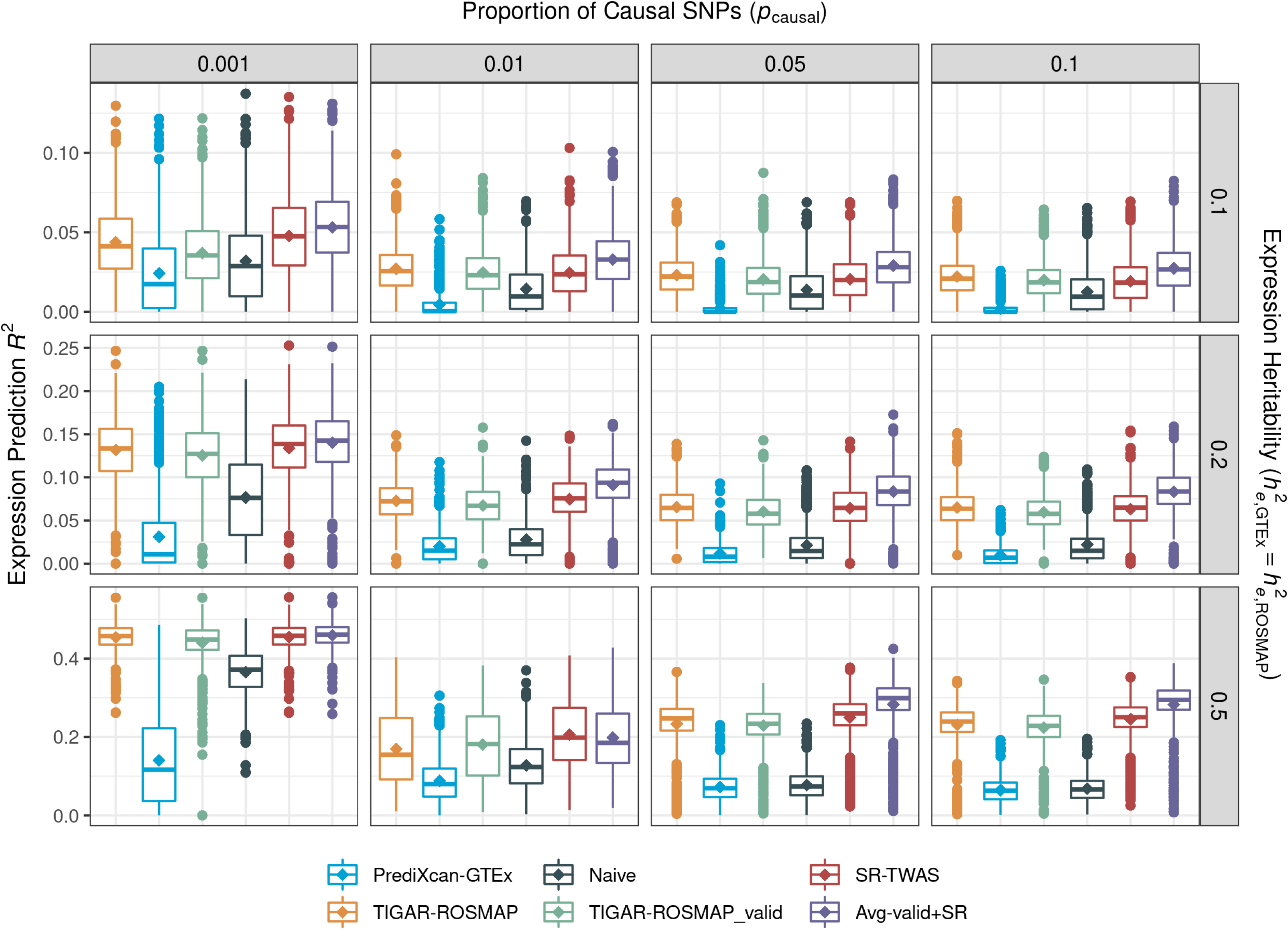
Boxplots of gene expression prediction *R*^2^ for simulations with varying proportion of true causal SNPs *p*_causal_ = (*0*. *001*, *0*. *01*, *0*. *05*, *0*. *1*) and true expression heritability *h_e_*^2^ = (*0*. *1*, *0*. *2*, *0*. *5*). Avg-valid+SR models obtained the highest test *R*^2^ for gene expression imputation across all 11 of 12 scenarios, while SR-TWAS performed best in the scenario with *p*_causal_ = 0.01 and *h_e_*^2^ = 0.5. This is because test samples are simulated under the same genetic architecture as the ROSMAP training cohort and the validation cohort, which only have ∼50% overlapped true causal SNPs as the GTEx training cohort.

As expected, model performance improved with increasing true expression heritability *h_e_*^2^ with the same training sample size. For all considered scenarios, the highest test *R*^2^ were obtained under a sparse causality model with *p*_causal_ = 0.001, where true causal SNP effect sizes would be relatively larger given the same *h_e_*^2^. The comparison of CV *R*^2^ and training *R*^2^ for Naïve and SR-TWAS approaches (Supplementary Figures 1-2) also showed that SR-TWAS outperformed the Naïve approach under all scenarios. Because the averaging step used to obtain Avg-valid+SR models does not include training and cross validation steps, no training *R*^2^ or CV *R*^2^ are obtained for comparison.

In order to assess TWAS power, phenotypes were simulated with a certain proportion of variance due to simulated gene expression (*h_p_*^2^). We considered a series of *h_p_*^2^ values in the range of (0.05, 0.875). The TWAS power comparison with three base models of the training and validation data (PrediXcan-GTEx, TIGAR-ROSMAP, and TIGAR-ROSMAP_valid), Naïve method, SR-TWAS, and Avg-valid+SR models were shown in **Fig. 2**, where the results were consistent with the test *R*^2^ comparison as in **Fig. 1**. The Avg-valid+SR approach performed best, followed by SR-TWAS, TIGAR-ROSMAP training base models, and TIGAR-ROSMAP_valid validation base models, while the Naïve method and the PrediXcan_GTEx training base models performed poorly in comparison. In the *p*_causal_ = 0.01 and *h_e_*^2^ = 0.5 scenario, SR-TWAS slightly outperformed Avg-valid+SR and had a noticeable advantage over the TIGAR-ROSMAP training base models and TIGAR-ROSMAP_valid validation models. The results showed the SR-TWAS approach indeed gained power by leveraging base models trained with multiple reference panels and by multiple statistical methods.

**Fig. 2.**
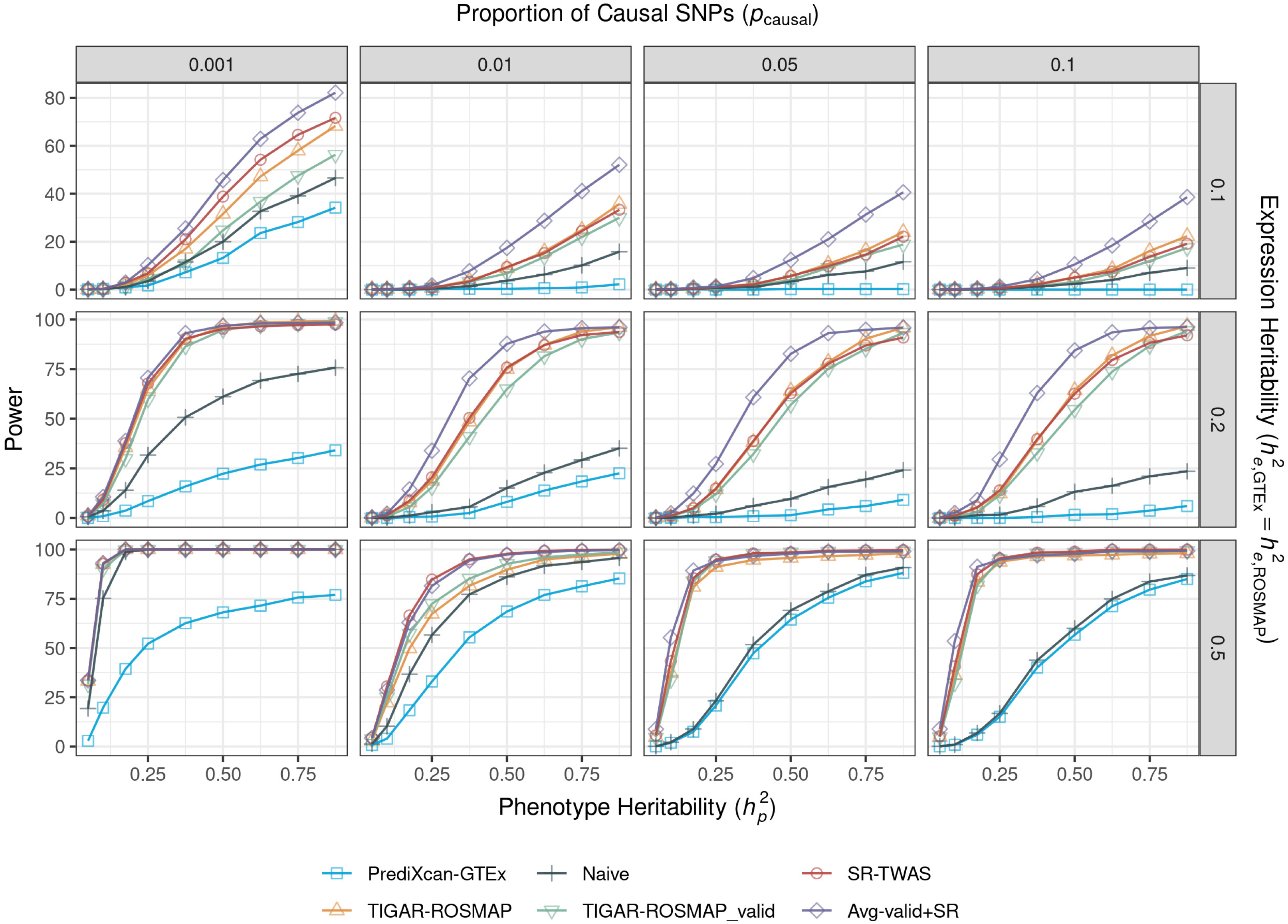
Power comparison for simulations with varying proportion of true causal SNPs *p*_causal_ = (0.001, 0.01, 0.05, 0.1), true expression heritability *h_e_*^2^ = (0.1, 0.2, 0.5), and phenotype heritability *h_p_*^2^ *∈* (0.05,0.875). Similar patterns were observed as shown in Fig. 1, where Avg-valid+SR models obtained the highest test *R*^2^ for gene expression imputation across all 11 of 12 scenarios and SR-TWAS models performed best in the scenario with *p*_causal_ = 0.01 and *h_e_*^2^ = 0.5.

Although desirable TWAS power ∼80% was only obtained in simulation scenarios with a relatively high *h_p_*^2^ that might be higher than the value in real studies, simulation power would increase along with increased test sample sizes. Because real GWAS test data would have a larger sample size than the 800 considered in our simulations, we expect desirable power for our SR-TWAS method in real studies.

#### Additional simulation studies

Additionally, we conducted similar simulation studies for two other settings, where samples from ROS/MAP and GTEx cohorts have the same set of true causal SNPs (i.e., the same genetic architecture), and (i) the expression heritability was the same for both ROS/MAP and GTEx V8 cohorts, or (ii) the expression heritability for GTEx V8 cohort was half that of the ROS/MAP cohort. The results of these settings were similar to that of the previously described setting. Avg-valid+SR still had the best performance, while SR-TWAS, TIGAR-ROSMAP training base models, and TIGAR-ROSMAP_valid validation base models outperformed the PrediXcan-GTEx training base models and Naïve models.

Comparisons of CV *R*^2^ and training *R*^2^ for Naïve and SR-TWAS approaches for these scenarios (Supplementary Figures 3, 4, 7, and 8) showed that SR-TWAS outperformed the Naïve approach under all scenarios. For all considered scenarios, again the highest test *R*^2^ was obtained under a sparse causality model with high expression heritability (Supplementary Figures 5 and 9). Power comparison results show that the Avg-valid+SR models obtained the highest power in most scenarios, while SR-TWAS, TIGAR-ROSMAP training base models, and TIGAR-ROSMAP_valid validation base models generally outperformed the PrediXcan-GTEx training base models and the Naïve models (Supplementary Figures 6 and 10). The SR-TWAS approach gained more power in the setting in which the expression heritability for GTEx V8 cohort was only half that of ROS/MAP (Supplementary Figure 10). SR-TWAS once again had the best performance in the *p*_causal_ = 0.01 and *h*^2^ = 0.5 scenario under both of these additional settings.

#### Model weights estimated by SR-TWAS

Plots of the weights (zeta values) of base models that were estimated by SR-TWAS in all three simulation settings and 12 scenarios (Supplementary Figure 11) showed that the SR-TWAS training consistently estimated higher weights for the TIGAR-ROSMAP training base models compared to the PrediXcan-GTEx training base models, with many models selecting only the TIGAR-ROSMAP training base model (ie, models in which the zeta value estimate for the TIGAR-ROSMAP training base model is 1 and the zeta value estimate for the PrediXcan-GTEx model is 0). This makes sense because the validation data were generated under the same model assumptions as the ROSMAP training data. In particular, when the GTEx training data were also generated under the same model as the validation data (orange bars in Supplemental Figure 11), zeta value estimates for the TIGAR-ROSMAP models are more evenly distributed in [0, 1] for scenarios with sparse causality model (p_causal = 0.001) and high expression heritability (he2 = 0.2, 0.5). When the GTEx training data were generated under a different setting where half of the true causal eQTL in the validation data were also causal in the ROSMAP training data (black bars in Supplemental Figure 11), the zeta value for the TIGAR-ROSMAP base model is more frequently estimated to be 1.

Even when both ROSMAP and GTEx training data were generated under the same model assumptions, the SR-TWAS method is still shown with gained power because the two base models trained by TIGAR and PrediXcan have complementary properties. In particular, PrediXcan uses a parametric penalized regression model with Elastic-Net penalty which is preferred for sparse genetic architecture of gene expression quantitative traits. Whereas TIGAR uses a nonparametric Bayesian Dirichlet process regression model which assumes an infinitesimal model for the underlying genetic architecture of gene expression quantitative traits.

As shown in previous studies^9^, PrediXcan will perform better when the true causal eQTL is sparse, while TIGAR will perform better as the true causal eQTL proportion increases. Our simulation studies showed that SR-TWAS had improved performance across various scenarios with (*p_causal_*= 0.001, 0.01, 0.05, 0.1; *h_e_*^2^ = 0.1, 0.2, 0.5; Supplementary Figure 5).

#### Type I error assessment

We also assessed type I error under the example scenario with *p*_causal_ = 0.1, *h_e_*^2^ = 0.1. Base model weights for TIGAR-ROSMAP, PrediXcan-GTEx, and TIGAR-ROSMAP-valid were permuted 10^6^ times. The TIGAR-ROSMAP and PrediXcan-GTEx training base models with permuted weights were then used to obtain SR-TWAS and Naïve models. The Avg-valid+SR was obtained by averaging the SR-TWAS models and TIGAR-ROSMAP_valid training base models with permuted weights. All models were then used to conduct gene-based association tests with a phenotype generated randomly from *N*(0, 1). As shown in Supplementary Table 2, all models controlled well for type I errors for significance thresholds (10^-4^, 10^-5^, 2.5 × 10^-6^, 10^-6^). The Quantile-Quantile (QQ) plots of the TWAS p-values in these null simulations are also shown in Supplementary Figure 12.

### Real Validation Studies

To compare the GReX prediction accuracy with real gene expression data, we considered three sets of base models that were trained by TIGAR with ROS samples (n=237, TIGAR_ROS_DLPFC) of dorsolateral prefrontal cortex (DLPFC) tissue, trained by TIGAR with GTEx V8 data of brain frontal cortex tissue (n=157, TIGAR_GTEx_BRNCTXB)^9^, and trained by PrediXcan with the same GTEx reference data of brain frontal cortex tissue (n=157, PrediXcan_GTEx_BRNCTXB)^7,8,18^. SR-TWAS (SR-TWAS_MAP_DLPFC) and Naïve (Naive_MAP_DLPFC) models were trained from these three sets of base models with respect to a validation dataset with half of the MAP samples (n=114, randomly selected) of DLPFC tissue. Valid gene expression imputation models trained by SR-TWAS and Naïve methods with 5-fold CV *R*^2^ > 0.5% in validation data were tested using the other half of the MAP samples (n=114) of DLPFC tissue.

By comparing test *R*^2^ obtained by SR-TWAS, Naïve, and three sets of base models in the test MAP samples (Supplementary Table 3), we showed that PrediXcan_GTEx_BRNCTXB^7,8,18^ had the highest median (0.070) and mean (0.113) test *R*^2^ but only for 867 valid gene expression imputation models, SR-TWAS had the second highest median (0.026) and mean (0.068) test *R*^2^ for 8425 valid genes expression imputation models, and Naïve model performed similarly to SR-TWAS but with a slightly lower median (0.025) and mean (0.065) test *R*^2^ and fewer valid genes expression imputation models (8360). By pair-wise comparison of test *R*^2^ for all genes with valid expression imputation models (Supplementary Figure 13), SR-TWAS (y-axis) performed noticeably better than the Naïve and three sets of base models (x-axis).

### Application TWAS of AD Dementia

#### Training expression imputation models of SMA tissue by SR-TWAS

We considered four sets of base models –– TIGAR and PrediXcan models trained with 465 ROS/MAP samples of DLPFC tissue (TIGAR_ROSMAP_DLPFC, PrediXcan_ROSMAP_DLPFC), TIGAR and PrediXcan models trained with 157 GTEx V8 samples of prefrontal cortex tissue (TIGAR_GTEx_BRNCTXB^9^, PrediXcan_GTEx_BRNCTXB ^7,8,18^). An additional 76 ROS/MAP samples of the supplementary motor area (SMA) brain tissue were used as the validation dataset to train SR-TWAS models and to calculate the 5-fold CV *R*^2^ that was used to select genes with valid imputation models. Plots of zeta weights estimated by SR-TWAS for each set of training base models were presented in Supplementary Figure 14. We observed that the weights of the TIGAR_GTEx_BRNCTXB^9^ training base models was estimated to be 1 more often than the other training base models. These results showed that the base models trained by TIGAR with the GTEx data of BRNCTXB tissue (TIGAR_GTEx_BRNCTXB^9^) contributed solely to SR-TWAS models for almost half of the genes, which was consistent with the numbers of valid gene expression imputation models as shown in Table 1.

**Table 1.** Comparison of CV *R*^2^ of SMA tissue for valid gene expression imputation models trained by training base models with ROS/MAP and GTEx V8 reference panels of DLPFC tissue, validation base models, SR-TWAS models, and Avg-valid+SR models.

| | Sample Size | Median CV $R^2$ | Mean CV $R^2$ | $N_{\text{genes}}$ |
| --- | --- | --- | --- | --- |
| PrediXcan_GTEx_BRNCTXB<sup>7,8,18a</sup> | 157 | 0.061 | 0.101 | 4563 |
| PrediXcan_ROSMAP_DLPFC<sup>a</sup> | 465 | - | - | 6532 |
| TIGAR_GTEx_BRNCTXB<sup>9a</sup> | 157 | 0.038 | 0.065 | 21921 |
| TIGAR_ROSMAP_DLPFC<sup>a</sup> | 465 | 0.017 | 0.049 | 11981 |
| PrediXcan_ROSMAP_SMA<sup>b</sup> | 76 | 0.031 | 0.053 | 23888 |
| TIGAR_ROSMAP_SMA | 76 | 0.064 | 0.077 | 32350 |
| SR-TWAS_ROSMAP_SMA | - | 0.072 | 0.090 | 20216 |
| Avg-valid+SR | - | - | - | 32238 |

*a.* Base training model used by SR-TWAS;
*b.* Base validation model.

PrediXcan and TIGAR models trained using the 76 ROS/MAP samples of SMA validation dataset (PrediXcan_ROSMAP_SMA, TIGAR_ROSMAP_SMA) were also included for comparison and were averaged with SR-TWAS models to obtain an additional set of models (Avg-valid+SR). Here, we compared Avg-valid+SR and SR-TWAS models to training base models, as well as validation base models trained by PrediXcan and TIGAR using the validation data of the target SMA brain tissue, to show the advantages of SR-TWAS for leveraging multiple regression models, multiple reference panels, and multiple tissues.

By comparing the CV *R*^2^ and numbers of genes with valid expression imputation models (**Table 1**), we found that gene expression imputation models trained by SR-TWAS for the SMA tissue (SR-TWAS_ROSMAP_SMA) had the highest median CV *R*^2^ (0.072) and second highest mean CV *R*^2^ (0.09) for ∼20K genes with valid expression imputation models. Although the PrediXcan_GTEx_BRNCTXB^7,8,18^ training base models had the third highest median CV *R*^2^ (0.061) and highest mean CV *R*^2^ (0.10), only 4563 genes had valid expression imputation models. These results showed that SR-TWAS obtained improved CV *R*^2^ in a real validation cohort of SMA tissue by leveraging multiple regression methods from two reference panels of multiple relevant tissues.

#### TWAS results of AD dementia

By using the eQTL weights obtained by training base models, validation base models, SR-TWAS models, and Avg-valid+SR models, we conducted TWAS with the summary-level data of the recent GWAS of AD dementia (n=∼762K)^19^. Since Avg-valid+SR models were shown to have the best performance in our simulation studies, here we focused on summarizing the results by Avg-valid+SR here. In particular, Avg-valid+SR identified a total of 89 significant TWAS risk genes of AD dementia with p-values < 2.5 × 10^-6^. Of these, 19 are known GWAS risk genes, 70 are within 1Mb of a known GWAS risk gene, and 14 have been previously observed as TWAS risk genes of AD dementia^20–25^ (Supplementary Table 4).

#### Validation of Significant TWAS Risk Genes of AD by PMR-Egger

In order to account for potential horizontal pleiotropy effects (genetic effects on phenotype that are not mediated by the considered GReX), we applied the PMR-Egger tool to the 89 significant TWAS risk genes of AD obtained by the Avg-valid+SR models. This analysis was performed using the same validation transcriptomic dataset and GWAS summary data as in the application TWAS of AD dementia. Of these 89 analyzed genes, 61 (68.5%) had a significant causal p-value by PMR-Egger after Bonferroni correction for multiple testing.

#### Independent significant TWAS risk genes of AD dementia

Because TWAS considers genotype data within a ±1Mb region of the test gene, nearby significant TWAS genes with overlapping test regions often have correlated GReX values and might not represent independent associations. We curated 6 independent TWAS risk genes of AD from the 61 significant genes that were validated by PMR-Egger (**Table 2**, **Fig. 3**) by selecting the most significant gene in a cluster of significant genes with overlapped test regions as the independent risk gene. We found that 1 of these independent risk genes, *HLA-DRA,* was a known GWAS^22,26^ risk gene and also previously observed as a TWAS risk gene^21^. The other 5 independent risk genes were near known GWAS risk genes^19,21,22,26–28^ and near previously observed TWAS risk genes^20,22^. Compared to the TWAS results using training base models (Supplementary Figure 15) and validation base models (**Fig. 3**), Avg-valid+SR models identified the greatest number of independent risk genes.

**Fig. 3.**
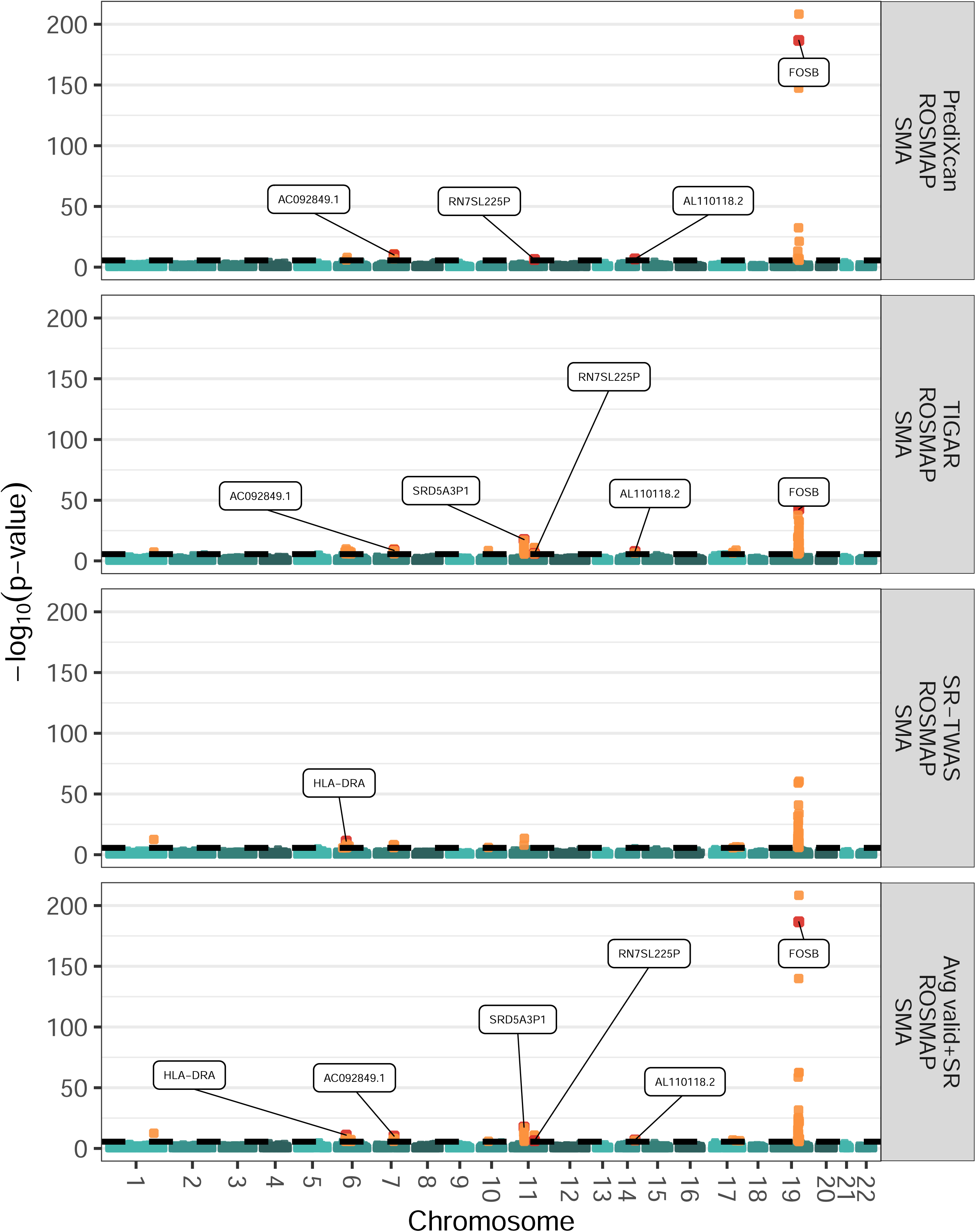
Manhattan plots of TWAS results by validation base models, SR-TWAS models, and Avg-valid+SR models of SMA tissue for studying AD dementia. Validation base models were trained by PrediXcan and TIGAR using the ROS/MAP validation data of SMA tissue. Avg-valid+SR models were obtained by averaging the SR-TWAS and these two sets of validation base models. A total of 89 (6 independent and significantly causal by PMR-Egger) TWAS risk genes were identified by the Avg-valid+SR models. Significant genes are shown in orange and significant genes discussed in the main text are labeled and shown in red.

**Table 2.** Independent TWAS risk genes of AD dementia identified by Avg-valid+SR models of SMA tissue. TWAS Zscores and p-values were presented here for the models of Avg-valid+SR, SR-TWAS, PrediXcan validation base models, and TIGAR validation base models. The signs of Zscores show the directions of the mediated genetic effects on AD dementia.

| Gene | CHR | Avg-valid+SR<br>ROSMAP<br>SMA |  | SR-TWAS<br>ROSMAP<br>SMA |  | PrediXcan<br>ROSMAP<br>SMA |  | TIGAR<br>ROSMAP<br>SMA |  |
| --- | --- | --- | --- | --- | --- | --- | --- | --- | --- |
|  |  | Zscore | P-value | Zscore | P-value | Zscore | P-value | Zscore | P-value |
| HLA-DRA<sup>ac</sup> | 6 | -6.81 | 1.00e-11 | -6.88 | 5.93e-12 | - | - | - | - |
| AC092849.1<sup>bd</sup> | 7 | 6.54 | 6.23e-11 | - | - | 6.48 | 9.02e-11 | 5.98 | 2.21e-09 |
| SRD5A3P1<sup>bd</sup> | 11 | 8.73 | 2.49e-18 | - | - | - | - | 8.73 | 2.49e-18 |
| RN7SL225P<sup>bd</sup> | 11 | 5.05 | 4.33e-07 | - | - | 5.03 | 5.02e-07 | 4.93 | 8.04e-07 |
| AL110118.2<sup>bd</sup> | 14 | -5.32 | 1.01e-07 | - | - | -5.25 | 1.52e-07 | -5.63 | 1.77e-08 |
| FOSB<sup>bd</sup> | 19 | 29.19 | 2.44e-187 | - | - | 29.20 | 1.87e-187 | 13.79 | 2.98e-43 |

*a.* Known GWAS risk gene of AD.
*b.* Gene within 1Mb of known GWAS risk gene of AD.
*c.* Previously observed TWAS risk gene of AD.
*d.* Gene within 1Mb of previously observed TWAS risk gene of AD.

#### Protein-Protein interaction network and enrichment analysis with risk genes of AD dementia

To investigate the underlying biological mechanisms of our identified TWAS risk genes of AD, we conducted protein-protein interaction network and enrichment analysis with our identified TWAS risk genes by the STRING^29^ tool (Methods). As shown in **Fig 5A**, we identified a major network consisting of 23 TWAS risk genes, including the well-known AD risk genes *TOMM40*^30^*, APOC1*^31^*, APOC2*^31^, and *TNF*^32^. Our identified TWAS risk genes are enriched with known risk genes for AD-related phenotypes such as family history of AD, lipoprotein measurements, mental or behavioral disorder biomarkers, inflammatory biomarker measurement, and beta-amyloid 1-42 measurement (**Fig 5B**).

### Application TWAS of PD

#### Training expression imputation models of brain substantia nigra tissue by SR-TWAS

We considered six sets of base models trained by TIGAR on six different tissues from GTEx V8 –– brain anterior cingulate cortex BA24 (BRNACC) (n=136), brain caudate basal ganglia (BRNCDT) (n=173), brain cortex (BRNCTXA) (n=184), brain nucleus accumbens basal ganglia (BRNNCC) (n=182), brain putamen basal ganglia (BRNPTM) (n=154), and whole blood (BLOOD) (n=574). With these six sets of training base models, an additional 101 GTEx samples of brain substantia nigra (BRNSNG) tissue were used as the validation data to train SR-TWAS models. We presented the plots of zeta weights of training base models that were estimated by SR-TWAS in Supplementary Figure 16. For all 6 sets of considered training base models, zeta weights were similarly distributed with 0’s for most genes and other values distributed over (0, 1). 5-fold CV *R^2^*s were calculated and used to select genes with valid expression imputation models for TWAS. PrediXcan and TIGAR models trained on the validation data of brain substantia nigra tissue (PrediXcan_GTEx_BRNSNG^7,8,18^, TIGAR_GTEx_BRNSNG^9^) were used to obtain average models (Avg-valid+SR) of these two sets of validation base models and SR-TWAS models.

By comparing the CV *R*^2^ and number of genes with valid expression imputation models (**Table 3**), we found that gene expression imputation models trained by SR-TWAS for the brain substantia nigra tissue (SR-TWAS_GTEx-BRNSNG^9^) had the highest median CV *R*^2^ (0.068) and highest mean CV *R*^2^ (0.094) for ∼23K genes with valid expression imputation models.

**Table 3.** Comparison of CV *R*^2^ of BRNSNG tissue for valid gene expression imputation models trained by training base models with GTEx V8 reference panel of multiple tissues, validation base models, SR-TWAS models, and Avg-valid+SR models.

| | Sample Size | Median CV $R^2$ | Mean CV $R^2$ | $N_{\text{genes}}$ |
| --- | --- | --- | --- | --- |
| TIGAR_GTEx_BRNACC<sup>9a</sup> | 136 | 0.040 | 0.062 | 23068 |
| TIGAR_GTEx_BRNCDT<sup>9a</sup> | 173 | 0.034 | 0.063 | 23362 |
| TIGAR_GTEx_BRNCTXA<sup>9a</sup> | 184 | 0.033 | 0.066 | 23379 |
| TIGAR_GTEx_BRNNCC<sup>9a</sup> | 182 | 0.033 | 0.060 | 23382 |
| TIGAR_GTEx_BRNPTM<sup>9a</sup> | 154 | 0.037 | 0.062 | 22481 |
| TIGAR_GTEx_BLOOD<sup>9a</sup> | 574 | 0.020 | 0.071 | 16682 |
| TIGAR_GTEx_BRNSNG<sup>9b</sup> | 101 | 0.050 | 0.068 | 23051 |
| PrediXcan_GTEx_BRNSNG<sup>7,8,18b</sup> | 101 | 0.052 | 0.080 | 2559 |
| SR-TWAS_GTEx_BRNSNG | - | 0.068 | 0.094 | 22913 |
| Avg-valid+SR | - | - | - | 21683 |

*a.* Base training model used by SR-TWAS;
*b.* Base validation model.

These results showed that SR-TWAS obtained improved CV *R*^2^ in a real validation cohort of substantia nigra tissue by leveraging multiple regression methods from two reference panels of multiple relevant tissues.

#### TWAS results of PD

We conducted TWAS using GWAS summary statistics by the recent GWAS of PD (n=∼33K cases, ∼18k UK Biobank proxi-cases, and ∼828K controls)^33^, using eQTL weights estimated by the above 6 sets of training base models of multiple tissues, validation base models of the BRNSNG tissue, SR-TWAS models, and Avg-valid+SR models (Supplementary Figure 17).

Here, we also focused on the results by using the Avg-valid+SR models (**Fig. 4**; **Table 4 ; Supplementary Table 5**), including a total of 60 significant TWAS risk genes of PD. Of these, 11 are known GWAS risk genes, 47 are within 1Mb of a known GWAS risk gene, and 11 have been previously observed as TWAS risk genes of PD.

**Fig. 4.**
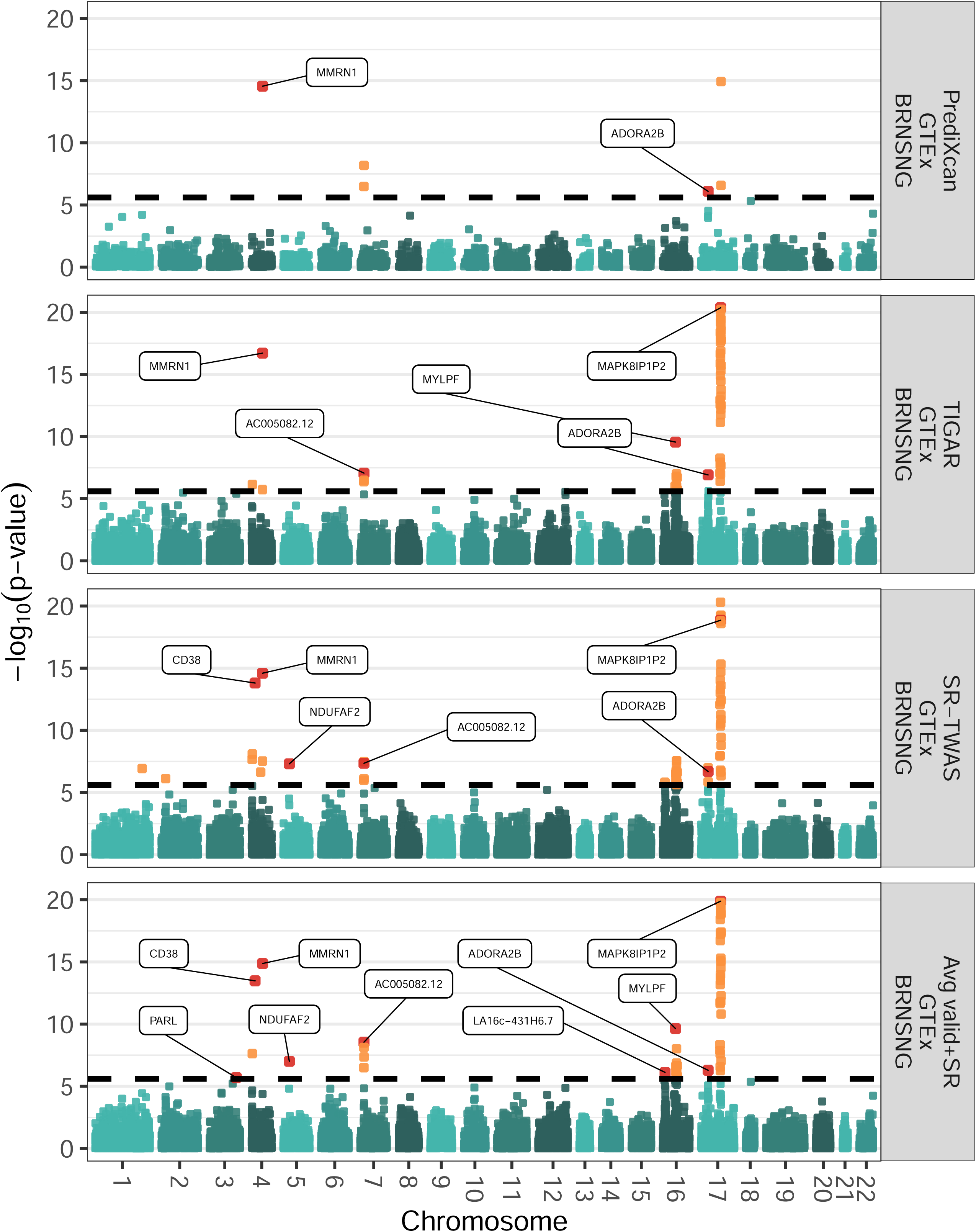
Manhattan plots of TWAS results by validation base models, SR-TWAS models, and Avg-valid+SR models of substantia nigra tissue for studying Parkinson’s Disease. Validation base models were trained by PrediXcan and TIGAR using the GTEx validation data of substantia nigra tissue. Avg-valid+SR models were obtained by averaging the SR-TWAS and these two sets of validation base models. A total of 60 (9 independent and significantly causal by PMR-Egger) significant TWAS risk genes were identified by the Avg-valid+SR models. Significant genes are shown in orange and significant genes discussed in the text are labeled and shown in red.

**Fig. 5.**
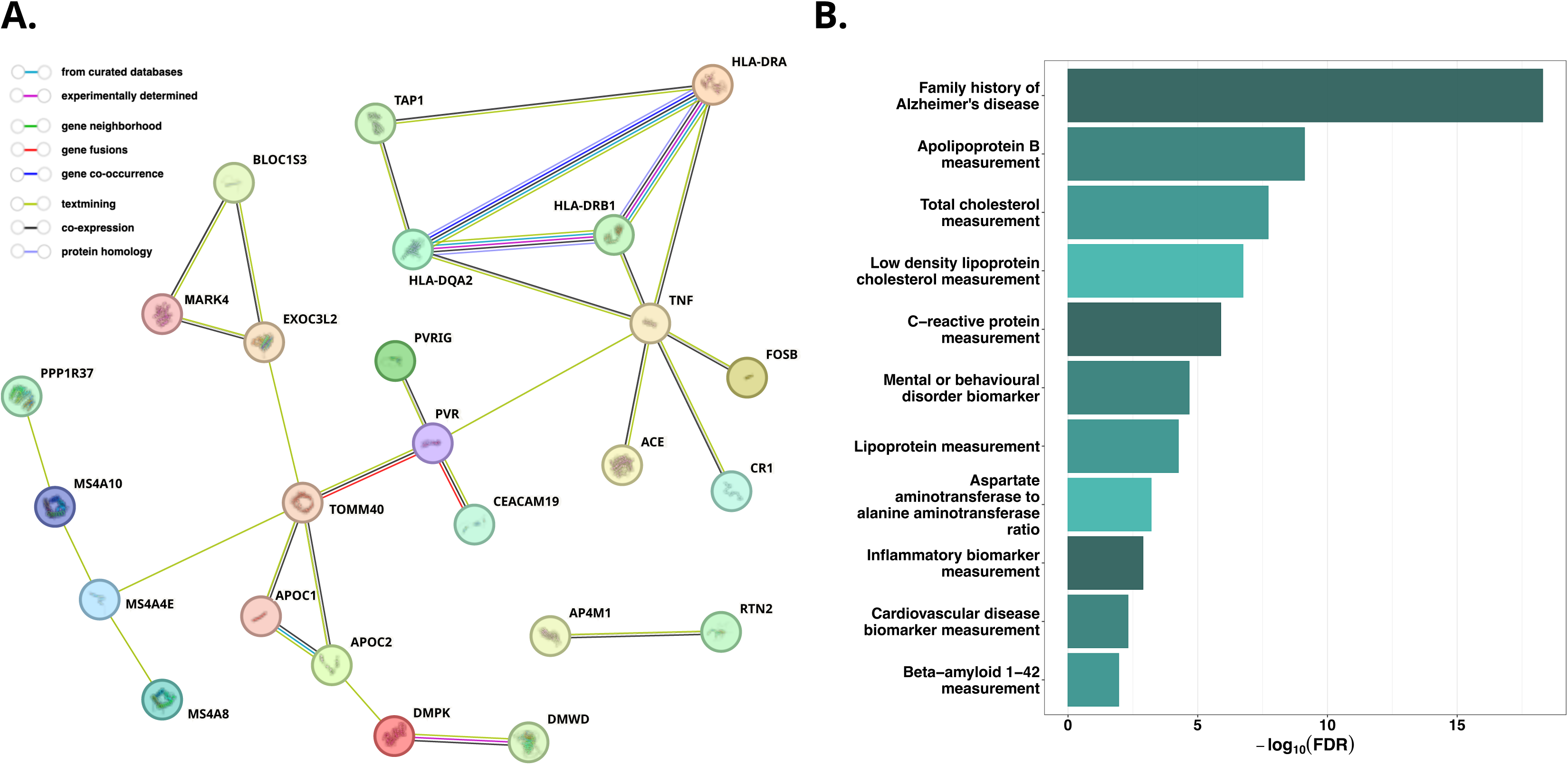
Protein-protein interaction network and enrichment analyses with TWAS risk genes of AD dementia by Avg-valid+SR models. A: A major network consisting of 23 TWAS risk genes is identified, including the well-known AD risk genes of *TOMM40, APOC1, APOC2*, and *TNF.* **B:** Enriched phenotypes include AD-related phenotypes such as family history of AD, lipoprotein measurements, mental or behavioral disorder biomarkers, inflammatory biomarker measurements, and beta-amyloid 1-42 measurement.

**Table 4.**
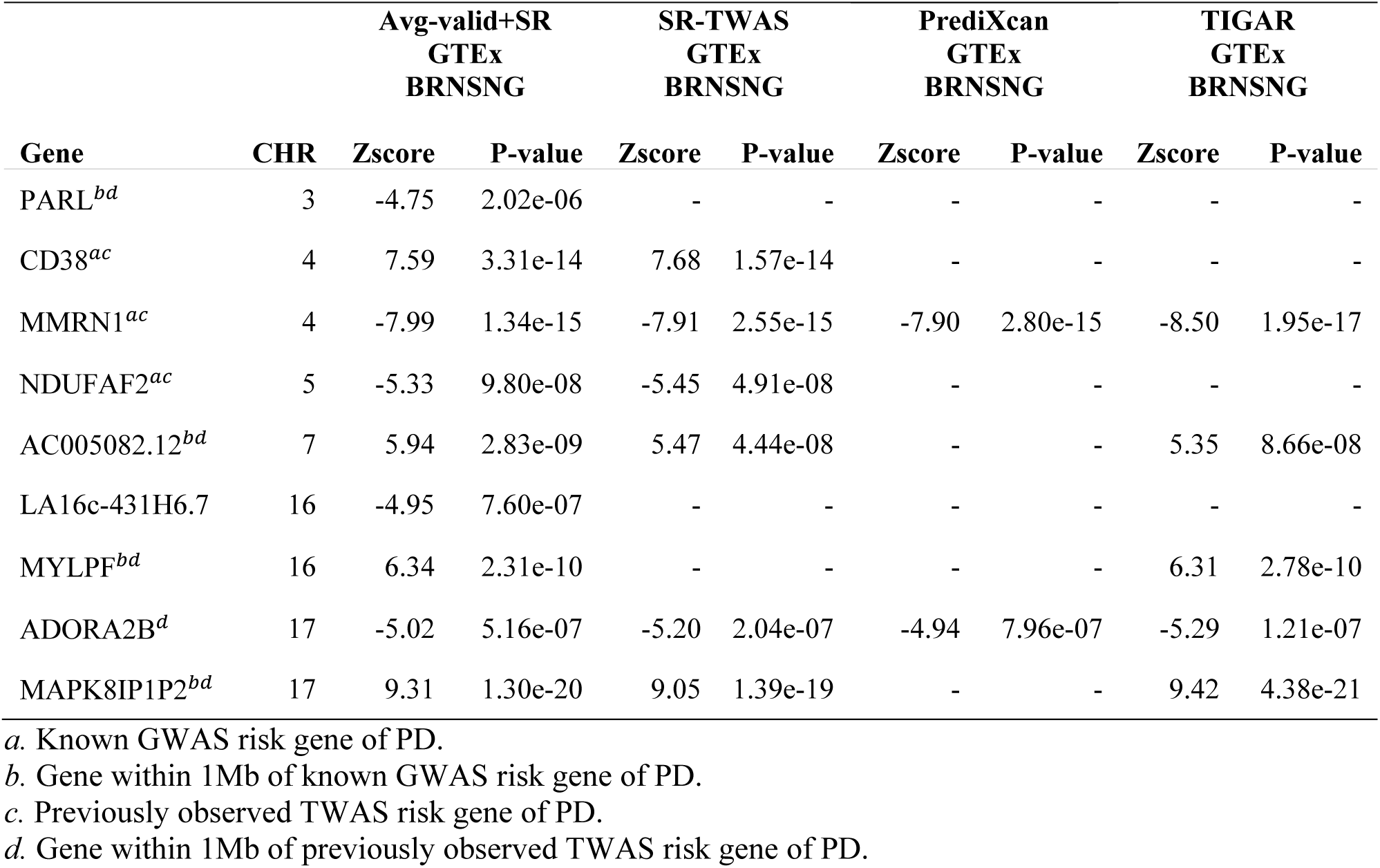
Independent TWAS risk genes of Parkinson’s disease identified by Avg-valid+SR models of SMA tissue. TWAS Zscores and p-values were presented here for the models of Avg-valid+SR, SR-TWAS, PrediXcan validation base models, and TIGAR validation base models. The signs of Zscores show the directions of the mediated genetic effects on AD dementia.

#### Validation of Significant TWAS Risk Genes of PD by PMR-Egger

We applied the PMR-Egger tool to the 60 significant TWAS risk genes of PD obtained by the Avg-valid+SR models. This analysis was performed using the same validation transcriptomic dataset and GWAS summary data as in the application TWAS of PD. Of these genes, 46 (76.6%) had a significant causal p-value by PMR-Egger after Bonferroni correction for multiple testing.

#### Independent Significant PD TWAS Risk Genes

Similarly, from these 46 replicated risk genes with significant causal p-values by PMR-Egger, we curated 9 independent TWAS risk genes of PD (**Fig. 4**; **Table 4**), including 6 novel TWAS risk genes (*LA16c-431H6.7*, *ADORA2B*, *AC005082.12*, *MAPK8IP1P2*, *MYLPF*, and *PARL*). Of these novel TWAS risk genes, four (*AC005082.12*, *MAPK8IP1P2*, *MYLPF*, and *PARL*) are near known GWAS risk genes (*GPNMB*^33,34^, *MAPT*^33^, *MCCC1*^33^, *SETD1A*^33^, *ZSWIM7*^35^). The other 3 previously observed TWAS risk genes were also known GWAS risk genes (*CD38*^34,35^, *MMRN1*^33,35^, and *NDUFAF2*^33^). Compared to the TWAS results using these six training base models (Supplementary Figure 17) and validation base models (**Fig. 4**), Avg-valid+SR models still identified the greatest number of independent risk genes that were validated by PMR-Egger.

#### Protein-Protein interaction network and enrichment analysis with risk genes of PD

Similarly, we conducted protein-protein interaction network and enrichment analysis with our identified TWAS risk genes of PD by the STRING^29^ tool (Methods). As shown in **Fig 6A**, we identified 4 networks with at least two connected genes, including a major one with 9 genes connected to the well-know PD risk gene *MAPT*^33^, and another network with 6 genes connected to gene *PRSS53,* a mapped PD risk gene in the GWAS Catalog^36^. Interestingly, our identified that TWAS risk genes of PD are enriched with known risk genes for mental traits such as anxiety, white matter microstructure measurement, handedness, and neuroticism measurement (**Fig 6B**).

**Fig. 6.**
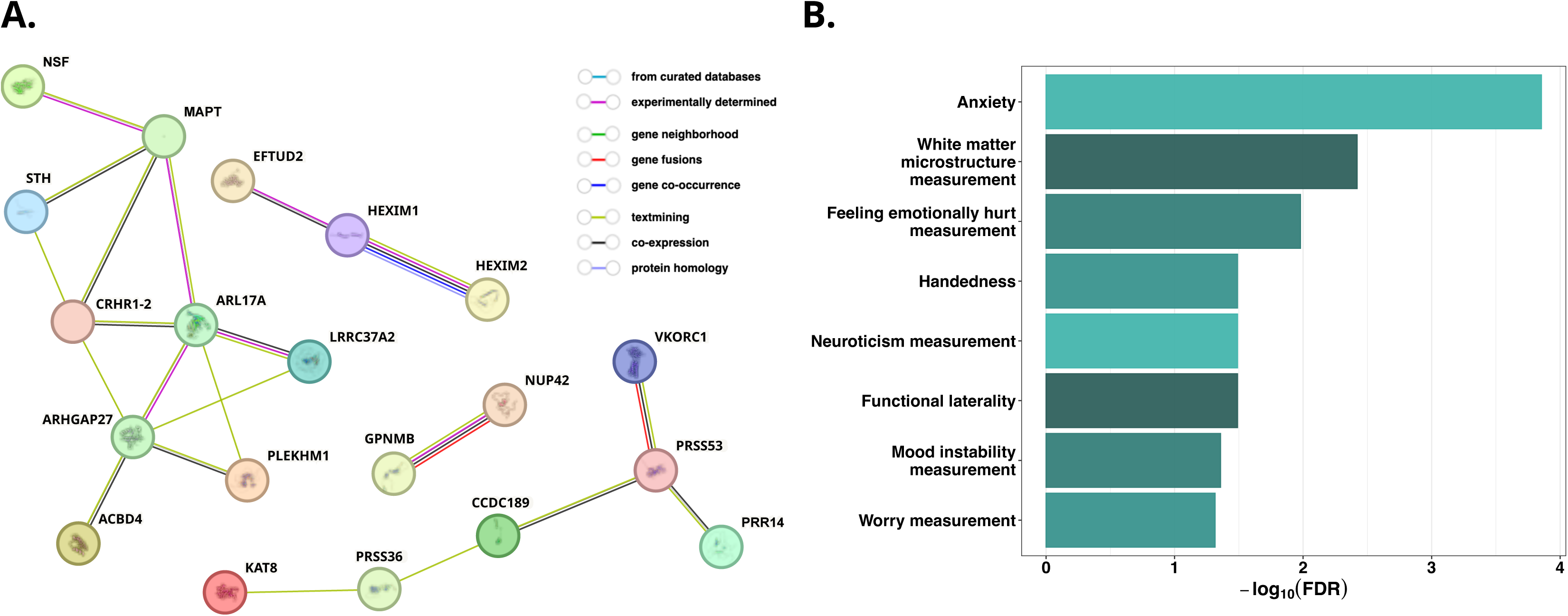
Protein-protein interaction network and enrichment analyses with TWAS risk genes of PD by Avg-valid+SR models. **A:** A total of 4 networks with at least two connected genes, including a major one with 9 genes connected through the well-known PD risk gene *MAPT*, and another one with 6 genes connected through *PRSS53*, which is a mapped PD risk gene in GWAS Catalog. **B:** Enriched phenotypes include mental traits such as anxiety, white matter microstructure measurement, handedness, and neuroticism measurement.

## Discussion

We present a novel TWAS tool (SR-TWAS) using the ensemble machine learning technique of stacked regression^15,16,37^ for leveraging multiple gene expression imputation models (i.e., base models) trained by different regression methods and/or using different transcriptomic reference panels of different tissue types. We also constructed a set of average models (Avg-valid+SR) of the SR-TWAS models and validation base models that were trained using the validation data. Different from existing methods such as UTMOST^12^ and SWAM^14^, SR-TWAS requires only summary-level base models, providing the flexibility of using publicly available base models.

With comprehensive simulation studies, we compared the Avg-valid+SR models to SR-TWAS models, the naïve approach of averaging all training base models, training base models, and validation base models. We showed that the Avg-valid+SR expression imputation models had the best prediction accuracy and led to the best TWAS power across 11 out of 12 considered scenarios, and that SR-TWAS models performed the best in the remaining scenario.

In the real data validation and application studies using ROS/MAP and GTEx V8 reference panels and GWAS summary data of Alzheimer’s disease (AD) dementia and Parkinson’s disease (PD), Avg-valid+SR models also outperformed base models trained using single reference panels and tissue types. Avg-valid+SR models identified a greater number of total independent risk genes that were replicated by PMR-Egger than any of the base training or validation models.

Besides known GWAS/TWAS risk genes that were identified by Avg-valid+SR models, we also found 5 novel independent TWAS risk genes for AD dementia and 6 novel independent TWAS risk genes for PD with known functions in respective disease pathology (Table 2; Table 4). Most of these novel findings are located within 1Mb region of previously known GWAS and TWAS risk genes. Additionally, we found interesting biological interpretations relevant to AD dementia and PD for our identified TWAS risk genes.

### Important findings by application TWAS of AD dementia

The results of the TWAS of AD include genes with known associations with AD dementia, or with relevant biological processes like immune response and regulation of AD-associated genes. Among these 6 curated independent significant genes by Avg-valid+SR models (**Table 2**), *HLA-DRA* is located in the major histocompatibility complex region that is expressed in glial cells^38^ and which has also been previously identified by eQTL analysis^22^. The other 5 independent significant TWAS AD risk genes of AD (*AC092849.1*, *AL110118.2*, *FOSB*, *RN7SL225P*, and *SRD5A3P1*) are within 1Mb of known GWAS risk genes of AD^19,21,22,26–28^ and previously observed TWAS risk genes^20,21^. *FOSB* was the most significant of a cluster of 38 identified significant TWAS risk genes with test region overlapped with the well-known GWAS risk gene *APOE*^19,21,22,28,39,40^ (**Fig. 3**). An alternatively-spliced product of the *FOSB* gene has been implicated in the regulation of gene expression and cognitive dysfunction in mouse models of AD^41^. *SRD5A3P* is located in the AD-associated *MS4A* gene cluster^27^, which contains multiple known GWAS risk genes^19,21–23^ as well as TWAS risk gene *MS4A2*^20^ of AD. The *MS4A* gene cluster is notable due its role in the regulation of soluble TREM2, which is encoded by the known AD risk gene *TREM2*, in cerebrospinal fluid in AD^27^.

### Important findings by application TWAS of PD

Similarly, results of the TWAS of PD include genes with known associations with PD, with related conditions, and with relevant biological processes like inflammation. Among these 9 curated independent significant genes (**Table 4**), *PARL* plays a role in regulating cellular processing of the mitochondrial kinase protein encoded by *PINK1*, mutations in which are a known cause recessively-inherited, early-onset PD^42^. *NDUFAF2* encodes for a component of mitochondrial complex I and loss of its functionality results in a rare mitochondrial encephalopathy with frequent substantia nigra pathology and motor symptoms^43^. *NDUFAF2* was also identified as a potential drug target in a Mendelian randomization study of potential drug targets for PD treatment^44^. A study of PD-associated *GPNMB* found that it is upregulated with lncRNA gene *AC005082.12*^45^.

Additionally, *ADORA2B* encodes adenosine receptor A2B which is an important cell receptor involved in numerous pathways and implicated in a broad variety of diseases including asthma, sepsis, inflammatory bowel disease, cancer, renal disease, diabetes, vascular diseases, and lung disease^46,47^. The immunomodulatory effects and role in inflammatory processes have made A2B a target for pharmacological therapeutics^46^and antagonists the similar adenosine receptor A2A were the first non-dopaminergic drug therapy for PD^48^. *MAPK8IP1P2* is a pseudogene near a known TWAS PD risk gene *LRRC37A2*^35^ (also identified by Avg-valid+SR models). *CD38* is involved in neurodegeneration, neuroinflammation, and aging^35,49^. *MMRN1* is a carrier protein for platelet factor V and lies *∼*84KB downstream of a well-established GWAS PD risk locus found in multiple populations^33^.

### Tool for implementing SR-TWAS

The SR-TWAS tool, including options for constructing SR-TWAS models, models by the Naïve method, and Avg-valid+SR models, is publicly-available on GitHub. The SR-TWAS tool implements user-friendly features, including accepting genotype data in standard VCF-format as input, enabling parallel computation, and using efficient computation strategies to reduce time and memory usage. The most computationally expensive part is to train all base models with different reference panels, which is subject to the regression method. For example, with training sample size n=465, PrediXcan (Elastic-Net) costs ∼1 CPU minute and TIGAR (DPR) costs ∼3 CPU minutes on average per gene. Publicly-available trained models can also be used as base models by the SR-TWAS tool. The process of training SR-TWAS models from base models and validation data is quite computational efficient. For example, with the ROS/MAP SMA tissue validation dataset (n=76) and four base models in our real studies SR-TWAS model training costs ∼15 CPU seconds per gene. With the GTEx substantia nigra tissue validation dataset (n=101) and six base models in our real studies SR-TWAS model training costs ∼103 CPU seconds per gene.

### Limitations

SR-TWAS still has its limitations. For example, SR-TWAS only considers cis-eQTL during model training, uses the standard two-stage TWAS, requires an additional validation dataset of the target tissue independent of those used for base model training^16^, and assumes samples of the reference panels and validation dataset are of the same ancestry^50^.

First, previous studies have illustrated the importance of considering both cis- and trans-eQTL in TWAS^51^, and a joint modeling of the gene expression imputation and the gene-based association test^52^ ^53^. The stacked regression technique used by SR-TWAS also applies to scenarios considering both cis- and trans-eQTL, when base models trained with both cis- and trans-eQTL are available.

Second, the standard two-stage TWAS framework (implemented by SR-TWAS) has limited power due to not considering the uncertainty of estimated eQTL weights and possible inflated false positives due to not considering potential horizontal pleiotropy (i.e., genetic effects on the phenotype of interest that are not mediated by GReX). Alternatively, a collaborative mixed model implemented by TWAS tools of CoMM^52^ and CoMM-S2^54^ that jointly model the reference transcriptomic and GWAS datasets (instead of two separate stages) is an effective approach to improve TWAS power by considering the uncertainty of estimated eQTL weights. The recently proposed PMR-Egger^55^ tool (for probabilistic mendelian randomization) can test the genetic effects mediated through the GReX term (equivalent to TWAS association tests) while controlling for horizontal pleiotropy could be used to validate the findings by SR-TWAS.

Third, in this study we only evaluated scenarios where reference samples used to train base models are of the same ancestry as the validation data. Although the method can be generalized to scenarios where base and validation data are of different ancestries, having at least one set of training base models with the same ancestry as the validation data would be a requirement for promising TWAS results. Evaluating the performance of SR-TWAS with base models of multiple ancestries is beyond the scope of this study, and is part of our ongoing study.

### Summary

Overall, the SR-TWAS tool provides a useful resource for researchers to take advantages of the publicly-available gene expression imputation models by using multiple regression methods (e.g., PrediXcan^7,8^, FUSION^17^, TIGAR^9^) and different reference panels of multiple tissue types (e.g., ROS/MAP^11^, GTEx V8^10^). In particular, the final trained gene expression imputation model by SR-TWAS will be with respect to the same tissue type as the validation data set. Because multiple base models would not only increase the robustness of the gene expression imputation model but also increase the total effective training sample size, SR-TWAS is expected to further increase TWAS power for studying complex human diseases. The approach of constructing average models of the SR-TWAS models and validation base models (Avg-valid+SR) provides a set of optimal gene expression imputation models that can leverage both training base models and validation base models to achieve the best TWAS performance.

## Acknowledgements

RP and JY are supported by National Institutes of Health (NIH/NIGMS) grant award R35GM138313. MPE was supported by NIH/NIGMS grant award R01GM117946 and NIH/NIA grant award RF1AG071170. ROS/MAP study data were provided by the Rush Alzheimer’s Disease Center, Rush University Medical Center, Chicago, IL. Data collection was supported through funding by NIA grants P30AG10161, R01AG15819, R01AG17917, R01AG30146, R01AG36836, R01AG56352, U01AG32984, U01AG46152, U01AG61356, the Illinois Department of Public Health, and the Translational Genomics Research Institute.

## Methods

### SR-TWAS using Stacked Regression

Stacked regression is a machine learning method for forming optimal linear combinations of different predictors to improve prediction accuracy^16^. The theoretical background for combining predictors rather than selecting a single best predictor is well-established and has been developed since the 1970s^16,56,57^. The “stacking” method of combining predictors originated in a 1992 paper^15^ by Wolpert, who described the concept as any scheme for feeding information from a set of cross-validated models to another before forming the final prediction in order to reduce prediction error^15^. The idea is further expanded with stacked regression, a specific framework for combining the initial predictors by weighted average with coefficient constraints to control for multicolinearity^16^.

In standard two-stage TWAS, we need to first fit a gene expression imputation model, which is assumed as a multivariable linear regression model, with quantitative gene expression levels *E_g_* for the target gene and tissue type as the response variable, and genotype matrix *G* of nearby/genome-wide SNPs as predictors,

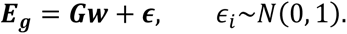

This gene expression imputation model can be trained per gene per tissue type, using a transcriptomic reference panel which profiles both transcriptomic and genetic data of the same training cohort. SNPs with non-zero effect sizes *w* are referred to as a broad sense of eQTL. The eQTL effect sizes *w* will be estimated from each trained model by different regression methods and/or using different reference data of multiple tissue types.

Assume there are a total of *K* base gene expression imputation models that are trained for the same target gene and tissue type, with 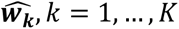 as the trained eQTL effect sizes per base model. Let **E***_vg_* denote the gene expression levels of the same target gene *g* and tissue type in the validation data, and **G***_v_* denote the genotype matrix of the same genetic predictors in the validation data. Then the predicted Genetically Regulated gene eXpression (GReX) of the validation samples are given by **G***_v_**ŵ**_k_*, by the *k*th base model. The stacked regression method^15,16^ will solve for a set of optimal model weights ζ*_1_*,…, ζ*_K_*, by maximizing the regression *R*^2^ between the profiled gene expression *E_vg_* and the weighted average GReX, 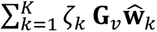 of *K* base models, ie, minimizing the following loss function of 1 *− R*^2^:

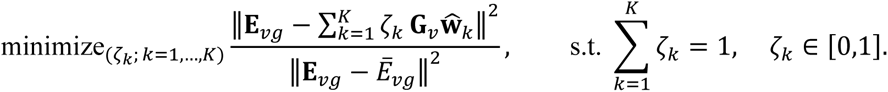

As a result, we will obtain a set of model weights *(_k_* for *k* = 1, …, *K* base models, and a set of eQTL effect sizes *w∼* given by the weighted average of the eQTL effect sizes of *K* base models, 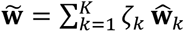 (Stage I). Then the final predicted GReX for test genotype data **G***_t_* is given by 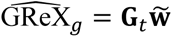, and *w∼* will be taken as variant weights in the gene-based association tests by SR-TWAS in Stage II.

Genes with 5-fold CV *R*^2^ > 0.5% in the validation dataset by SR-TWAS are considered as having a valid imputation model and will be tested in Stage II. That is, the validation dataset will be randomly split into 5 folds. For each fold of data, SR-TWAS model will be trained using the other 4-fold data and then use to calculate prediction *R*^2^ with the current fold. The average prediction *R*^2^ across all 5 folds of data is considered as the 5-fold CV *R*^2^. Here, we use a more liberal threshold (0.005) than the threshold 0.01 used by previous studies ^17,58,59^ to allow more genes to be tested in follow-up TWAS. Because the follow-up gene-based association Z-score test statistic is essentially a weighted average of single variant GWAS Z-score statistics with variant weights provided by the eQTL effect sizes^9^, poorly estimated eQTL weights would only reduce power but will not increase false positive rate under null hypothesis.

### Naïve Method

In this paper, we compared SR-TWAS to a Naïve approach which just takes the average of base models as the trained gene expression imputation model, that is, takes 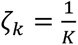, *k* = 1, …, *K*. Using a validation dataset, we can still evaluate the validation *R*^2^ which can be used to select valid genes with validation *R*^2^ > 0.5%.

### Avg-valid+SR Models

We further constructed average models of the SR-TWAS models and validation base models trained using the validation dataset, which are referred to as Avg-valid+SR models. Because SR-TWAS and validation base models are averaged directly, training *R*^2^ and CV *R*^2^ are not obtained for Avg-valid+SR models. We compared the Avg-valid+SR models to the SR-TWAS models and validation base models in both simulation and real studies.

### SR-TWAS Tool Framework

SR-TWAS tool was designed to be compatible with the TIGAR-V2 tool framework^9^; it accepts models trained by TIGAR-V2 as input, imports utility functions from TIGAR-V2, and outputs model files which can be used as input for TIGAR-V2 GReX prediction and summary-level TWAS. Much of the structure of the SR-TWAS code was derived from existing TIGAR-V2 scripts and it shares dependencies on TABIX^60^ and the Python libraries of numpy^61,62^, pandas^61^, scipy^63^, statsmodels^64^, and scikit-learn^65,66^.

The SR-TWAS script utilizes scikit-learn’s consistent, extensible interfaces for defining estimators and predictors and for initializing objects^66^. The script trains a stacked regression model using a modified version of scikit-learn’s StackingRegressor class, which trains a final estimator from cross-validated predictions from base estimators fitted on the full design matrix. The script defines two custom classes to be used as input for the stacking regressor object: a base estimator class (WeightEstimator) which converts trained GReX prediction models into scikit-learn-compatible estimator objects and a final estimator class (ZetasEstimator) which obtains the values of *ζ_1_*,…, *ζ_K_* that minimize the loss function under the constraints *ζ_k_ ≥* 0 and 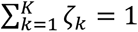^16^.

During the stacked regression, SNP minor allele frequencies and effect sizes for the specified target are first read from each of the *K* user-specified weight files. The SNPs are then matched to SNPs in the validation genotype data and filtered to exclude effect sizes of SNPs for which the difference between the MAF of the genotype data and the MAF from the corresponding weight file exceeds a user-specified MAF difference threshold. The effect sizes from each weight file are used to initialize *K* separate instances of the WeightEstimator class. These *K* WeightEstimator objects are used as base estimators and fit on genotype and expression data from the validation data.

A separate script allows users to specify one or more base models trained on the validation data to average with the SR-TWAS model produced in the previous step. The tool will read SNP effect sizes for the specified target from each of these models and output an Avg-valid+SR model averaged from the validation data base model(s) and the model obtained by stacked regression.

Only SR-TWAS models trained from *K* = 2, 4,6 base models are presented in this paper. The code was designed to accept any *K ≥* 2, and while the stacked regression script has been primarily tested using *K* = 2, 4, 6 base models, preliminary testing with dummy weight files confirms it can train stacked regression models from *K* > 6 base models.

### ROS/MAP Reference Panel

The Religious Orders Study (ROS) and Rush Memory and Aging Project (MAP) are two ongoing longitudinal, epidemiologic clinical-pathologic cohort studies of aging and Alzheimer’s disease collectively referred to as ROS/MAP^11^. ROS enrolls Catholic nuns, priests, and brothers from religious groups across the United States, primarily from communal living settings^11^. While the similar adult lifestyle of participants allows for more control of potential confounders such as education and socioeconomic status, it simultaneously limits the ability to study such variables^11^. MAP was designed to complement and extend studies like ROS by including subjects from a wider range of life experiences, socioeconomic status, and educational attainment and recruits participants primarily from retirement communities in the Chicago area, but also subsidized housing, retirement homes, and through organizations serving minorities and low-income elderly^11^. All participants in both studies are without known dementia and agree to annual clinical evaluations and brain donation upon death^11^. Similarity in study design and data collection procedures allows the ROS and MAP datasets to be merged for use in joint analyses^11,67^.

Quality-controlled ROS/MAP WGS data for European subjects^67^ were used for both the real data application and simulation studies. Transcriptomic data of ROS/MAP samples of brain PFC were profiled by RNA-sequencing (RNA-seq). Gene expression data of Transcripts Per Million (TPM) per sample were provided by Rush Alzheimer’s Disease Center. Genes with > 0.1 TPM in *≥* 10 samples were considered. Raw gene expression data (TPM) were then log2 transformed and adjusted for age at death, sex, postmortem interval, study (ROS or MAP), batch effects, RNA integrity number scores, cell type proportions (with respect to oligodendrocytes, astrocytes, microglia, neurons), top five genotype principal components, and top probabilistic estimation of expression residuals (PEER) factors^68^ by linear regression models. SNPs with minor allele frequency (MAF) > 1%, Hardy-Weinberg p-value > 10^-5^ were analyzed. For each gene, cis-SNPs within the 1Mb of the flanking 5’ and 3’ ends were used in the imputation models as predictors.

### GTEx V8 Reference Panel

The Genotype-Tissue Expression (GTEx) project V8 profiles both whole genome sequencing (WGS) genotype data and RNA-seq transcriptomic data of 54 human tissues^10^. The fully processed, filtered, and normalized transcriptomic data used in the GTEx eQTL analysis were downloaded from the GTEx portal and used in this study. For each tissue, samples with <10 million mapped RNA-seq reads were excluded. For samples with replicates, the replicate with the greatest number of reads was selected. Gene read counts from each sample were normalized using size factors calculated by DESeq2 and log-transformed with an offset of 1. Genes with log-transformed value > 1 in > 10% samples were considered. The resulting gene expression values were centered with mean 0 and standardized with standard deviation 1. The resulting matrix was then hierarchically clustered (based on average and cosine distance), and a chi2 p-value was calculated based on Mahalanobis distance. Clusters with ≥60% samples with Bonferroni-corrected p-values <0.05 were marked as outliers, and their samples were excluded. Genetic variants with missing rate < 20%, minor allele frequency > 0.01, and Hardy-Weinberg equilibrium p-value > 10^-5^ were considered for fitting the gene expression prediction models.

The fully processed, filtered, and normalized transcriptomic data were adjusted for top five genotype principal components, top probabilistic estimation of expression residuals (PEER) factors^68^, sequencing protocol (PCR-based or PCR-free), sequencing platform (Illumina HiSeq 2000 or HiSeq X), and sex, as suggested by the GTEx eQTL data analysis guidelines^10^. The number of top PEER factors used to adjust the gene expression traits depends on the sample size (n) in the reference transcriptomic data cohort –– 15 factors for n<150, 30 factors for 150≤n<250, 45 factors for 250≤ n<350, and 60 factors for n≥350. Only samples with complete data of these covariates were included in the analyses. Adjusted gene expression quantitative traits were then taken as response variables in the gene expression prediction model. For each gene, cis-SNPs within the 1Mb of the flanking 5’ and 3’ ends were used in the imputation models as predictors.

### Simulation Study Design

We conducted in depth simulation studies under various scenarios to assess the performance of SR-TWAS, Avg-valid+SR, a Naïve method, and training base models by PrediXcan and TIGAR. We used the real genotype data of gene *ABCA7* from ROS/MAP and GTEx V8 to simulate gene expression and phenotypes. We considered three different settings: (i) Samples from ROS/MAP and GTEx cohorts have the same set of true causal SNPs (i.e., the same genetic architecture). The expression heritability was the same for both ROS/MAP and GTEx V8 cohorts. (ii) Samples from ROS/MAP and GTEx cohorts have the same set of true causal SNPs (i.e., the same genetic architecture). The expression heritability for GTEx V8 cohort is only half of the one for ROS/MAP. (iii) Samples from the ROS/MAP cohort were simulated with the same causal SNPs (i.e., eQTL), while samples from the GTEx V8 cohort were simulated with true causal SNPs that were 50% overlapped with the ones for ROS/MAP. The expression heritability was the same for both ROS/MAP and GTEx V8 cohorts.

Under each setting, we considered multiple scenarios with varying proportions of causal SNPs (*p_causal_* = (0.001, 0.01, 0.05, 0.1)) and gene expression heritability (i.e., the proportion of gene expression variation due to genetics, *h*^2^ = (0.1, 0.2, 0.5)). We randomly selected n=465 training samples with WGS genotype data from ROS/MAP and GTEx V8, respectively. We randomly selected n=400 and n=800 samples with WGS genotype data from ROS/MAP as our validation and test cohorts, respectively. We considered a series of *h*^2^ values, the proportion of phenotype variance due to simulated gene expression, in the range of (0.05, 0.875).

For each scenario, gene expression *E_i_* for the *itℎ* simulation iteration is generated using the following formula

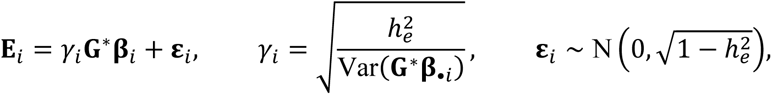

where **G***^∗^* denotes the genotype matrix of *N*_causal_ randomly chosen true causal SNPs for all samples, effect size vector *JJ_i_* was generated from *N*(0, *I*), and *y_i_* is a scale factor chosen to ensure the targeted *h_e_*^2^ value. The phenotype vector *Y_i_* for the *i*th simulation iteration was generated using the following formula

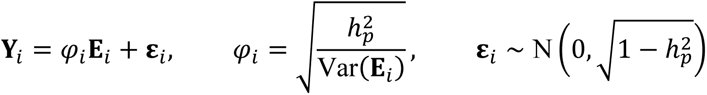

where **E***_i_*is the simulated gene expression, and *φ_i_* is a scale factor to ensure the targeted *h_p_*^2^ value.

Two base models per gene were trained by PrediXcan with the GTEx training samples (n=465) (PrediXcan-GTEx), and by TIGAR with the ROS/MAP training samples (n=465) (TIGAR-ROSMAP). SR-TWAS and Naïve models were then obtained by using these trained base models. Validation data (*n* = 400) were used to train SR-TWAS models and filter out gene expression imputation models with 5-fold cross-validation *R*^2^ < 0.5% in the validation cohort for both SR-TWAS and Naïve models. A validation data base model was trained by TIGAR on the validation data (TIGAR-ROSMAP_valid) to compare results with that of the ensemble models and to obtain a model from the average of the the validation data base models and SR-TWAS (Avg-valid+SR). Test data (n=800) were used for assessing GReX prediction performance and TWAS power. Each causal simulation scenario was repeated for 1,000 times. We compared the performance by SR-TWAS, Avg-valid+SR, the Naïve method, training base models, and validation base models with respect to prediction imputation *R*^2^ in the test data and the power of TWASs.

The predicted 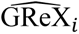 by each trained gene expression imputation model was used to calculate expression prediction *R*^2^, which is equivalent to the regression *R*^2^ between profiled and predicted gene expression, given by

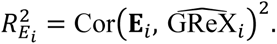

The power will be given by the proportion of simulation iterations that have TWAS p-value < 2.5 × 10^6^ out of a total of 1,000 simulation iterations.

### Protein-Protein Interaction Network and Enrichment Analysis

STRING (version 12.0)^29^ is a bioinformatics web tool that provides information on protein-protein interactions and networks, as well as functional characterization of genes and proteins. The tool integrates different types of evidence from public databases, such as genomic context, high-throughput experiments, and previous knowledge from other databases, to generate reliable predictions of protein interactions and build networks and pathways. Provided with a list of gene names, STRING will construct networks based on the protein-protein interactions of the corresponding proteins, as well as identify phenotypes that have risk genes enriched in the provided list. Proteins corresponding to provided genes are considered nodes in the protein-protein interaction network. Protein-protein edges represent the predicted functional associations, and their color denotes one of seven different evidence categories –– computational interaction predictions from co-expression, “text-mining” of scientific literature, databases of interaction experiments (biochemical/genetic data), known protein complexes or pathways from curated resources, gene co-occurrence, gene fusion, and gene neighborhood. Gene co-occurrence, fusion, and neighborhood represent association predictions based on whole-genome comparisons. Interactions from these resources are critically assessed, scored, and subsequently automatically transferred to less well-studied organisms using hierarchical orthology information^29^.

Particularly, the “text-mining” channel is the result of parsing full-text articles from the PMC Open Access Subset (up to April 2022), PubMed abstracts (up to August 2022), as well as summary texts from OMIM^69^ and Saccharomyces genome database^70^ entry descriptions. These texts are all parsed for co-mentions of protein pairs and assessed against the frequencies of all separate mentions of the respective proteins. An improved deep learning-based relation extraction text mining model was used by STRING v12^29^. The “text-mining” channel significantly increases the number of protein–protein interactions.

## Data availability

All ROS/MAP data analyzed in this study are de-identified and available to any qualified investigator with application through the Rush Alzheimer’s Disease Center Research Resource Sharing Hub, https://www.radc.rush.edu, which has descriptions of the studies and available data. GTEx V8 data are available from dbGaP with accession phs000424.v8.p2, and GTEx Portal https://www.gtexportal.org/home/. TIGAR DPR base models trained from GTEx V8 are available at Synapse https://www.synapse.org/TIGAR_V2_Resource_GTExV8. PrediXcan Elastic-Net base models trained from GTEx V8 are available from https://predictdb.org/. GWAS summary data of AD are available from https://ctg.cncr.nl/software/summary_statistics, and GWAS summary data of PD are available from https://bit.ly/2ofzGrk. TIGAR DPR and PrediXcan Elastic-Net base models of ROS/MAP tissues (DLPFC, SMA), SR-TWAS and Avg-valid+SR models trained from ROS/MAP SMA tissue and GTEx brain substantia nigra tissue in this study, and all TWAS summary statistics generated in this study are freely available from SYNAPSE https://doi.org/10.7303/syn53437281.

## Code availability

The SR-TWAS tool, including the Naïve and Avg-valid+SR methods, is publicly-available on GitHub, https://github.com/yanglab-emory/SR-TWAS. Code for replicating analyses described in this paper is available at https://github.com/rndparr/SR-TWAS_analysis.

