## Supplemental Figures and Tables for "SR-TWAS: Leveraging Multiple Reference Panels to Improve TWAS Power by Ensemble Machine Learning"

### Supplemental Tables

**Supplementary Table 1: Disease status of GTEx and ROS/MAP subjects**

| TWAS | Use | Dataset | N | Normal | Disease* | Missing |
|---|---|---|---|---|---|---|
| AD | Train | GTEx | 158 | 150 (95%) | 7 (4%) | 1 (1%) |
| AD | Train | ROS/MAP | 465 | 263 (57%) | 185 (40%) | 17 (4%) |
| AD | Validation | ROS/MAP | 76 | 57 (75%) | 19 (25%) | 0 (0%) |
| PD | Train | GTEx | 640 | 554 (87%) | 3 (0.4%) | 83 (13%) |
| PD | Validation | GTEx | 101 | 94 (93%) | 1 (1%) | 6 (6%) |

∗ Disease status of AD or other types dementia for samples used in AD-TWAS; Disease status of PD for samples used in PD-TWAS.

**Supplementary Table 2: Assessment of type 1 error that is the proportion of significant tests among $10^6$ null simulations.**

| Significance Threshold | TIGAR ROSMAP | PrediXcan GTEx | TIGAR ROSMAP valid | Naive ROSMAP valid | SR-TWAS ROSMAP valid | Avg-valid+SR ROSMAP valid |
|---|---|---|---|---|---|---|
| $1.00 \times 10^{-4}$ | $9.60 \times 10^{-5}$ | $9.30 \times 10^{-5}$ | $9.90 \times 10^{-5}$ | $8.60 \times 10^{-5}$ | $7.70 \times 10^{-5}$ | $9.30 \times 10^{-5}$ |
| $1.00 \times 10^{-5}$ | $8.00 \times 10^{-6}$ | $1.10 \times 10^{-5}$ | $1.30 \times 10^{-5}$ | $9.00 \times 10^{-6}$ | $9.00 \times 10^{-6}$ | $1.20 \times 10^{-5}$ |
| $2.50 \times 10^{-6}$ | $1.00 \times 10^{-6}$ | $4.00 \times 10^{-6}$ | $6.00 \times 10^{-6}$ | $3.00 \times 10^{-6}$ | $3.00 \times 10^{-6}$ | $4.00 \times 10^{-6}$ |
| $1.00 \times 10^{-6}$ | $1.00 \times 10^{-6}$ | $1.00 \times 10^{-6}$ | $3.00 \times 10^{-6}$ | $1.00 \times 10^{-6}$ | $1.00 \times 10^{-6}$ | $1.00 \times 10^{-6}$ |

**Supplementary Table 3: Test $R^2$ in real validation studies for genes with CV $R^2 > 0.5\%$.**

| Model | Panel | Tissue | Median $R^2$ | Mean $R^2$ | $N_{\text{genes}}$ |
|---|---|---|---|---|---|
| PrediXcan | GTEx | BRNCTXB | 0.07080 | 0.11395 | 867 |
| TIGAR | GTEx | BRNCTXB | 0.02305 | 0.05837 | 2405 |
| TIGAR | ROS | DLPFC | 0.02358 | 0.06446 | 7913 |
| Naive | MAP | DLPFC | 0.02576 | 0.06515 | 8360 |
| SR-TWAS | MAP | DLPFC | 0.02606 | 0.06820 | 8425 |

**Supplementary Table 4: Significant TWAS risk genes of AD dementia identified by Avg-valid+SR models for the ROSMAP validation data of SMA tissue. TWAS Zscores and p-values were presented here for the models of Avg-valid+SR, SR-TWAS, PrediXcan base validation models, and TIGAR base validation models. The signs of Zscores show the directions of the mediated genetic effects on AD dementia.**

| | | Avg-valid+SR ROSMAP-SMA | | SR-TWAS ROSMAP-SMA | | PrediXcan ROSMAP-SMA | | TIGAR ROSMAP-SMA | |
|---|---|---|---|---|---|---|---|---|---|
| Gene | Locus | Zscore | Pvalue | Zscore | Pvalue | Zscore | Pvalue | Zscore | Pvalue |
| CR1[ac] | 1:207496147-207641765 | -7.3 | 2.88e-13 | -7.3 | 2.91e-13 | - | - | - | - |
| TNF[bd] | 6:31575565-31578336 | -5.45 | 4.99e-08 | - | - | - | - | -5.64 | 1.67e-08 |
| MSH5-SAPCD1[bd] | 6:31740020-31764851 | 4.89 | 9.91e-07 | - | - | - | - | 4.89 | 9.91e-07 |
| TSBP1-AS1[bd] | 6:32254640-32407763 | -5.22 | 1.79e-07 | -5.22 | 1.79e-07 | - | - | - | - |
| HLA-DRA[ac] | 6:32439878-32445046 | -6.81 | 1.00e-11 | -6.88 | 5.93e-12 | - | - | - | - |
| HLA-DRB1[ac] | 6:32578769-32589848 | -4.84 | 1.28e-06 | -5.21 | 1.87e-07 | - | - | - | - |
| HLA-DQA2[bd] | 6:32741391-32747198 | 4.96 | 6.88e-07 | 5.98 | 2.26e-09 | 4.87 | 1.13e-06 | 5.18 | 2.26e-07 |
| TAP1[bd] | 6:32845209-32853978 | -5 | 5.61e-07 | -4.99 | 6.02e-07 | - | - | - | - |
| AL645941.1[bd] | 6:32970232-32970886 | -5.87 | 4.23e-09 | - | - | - | - | -5.87 | 4.23e-09 |
| KIFC1[bd] | 6:33391823-33409896 | -5.22 | 1.81e-07 | -4.81 | 1.49e-06 | - | - | - | - |
| SYNGAP1-AS1[bd] | 6:33437363-33454453 | 4.83 | 1.33e-06 | - | - | - | - | 4.83 | 1.33e-06 |
| ZBTB9[bd] | 6:33453970-33457544 | -4.93 | 8.03e-07 | - | - | - | - | -4.93 | 8.03e-07 |
| GGNBP1[bd] | 6:33540046-33589026 | 5.76 | 8.28e-09 | - | - | 5.75 | 8.80e-09 | 5.27 | 1.40e-07 |
| ANKRD66[bd] | 6:46746917-46759506 | 4.78 | 1.72e-06 | - | - | - | - | 4.78 | 1.72e-06 |
| AL355353.1[bd] | 6:47489067-47489170 | 5.45 | 5.02e-08 | - | - | - | - | 5.45 | 5.02e-08 |
| ATP5MF-PTCD1[bd] | 7:99419749-99466197 | 5.29 | 1.25e-07 | - | - | - | - | 5.29 | 1.25e-07 |
| ZSCAN25[bd] | 7:99616946-99632408 | 5.68 | 1.36e-08 | 5.77 | 8.14e-09 | - | - | - | - |
| OR2AE1[bd] | 7:99875987-99877057 | -5.37 | 7.85e-08 | - | - | - | - | -5.37 | 7.85e-08 |
| AZGP1P1[bd] | 7:99980762-99987535 | -5.25 | 1.54e-07 | - | - | - | - | -5.25 | 1.54e-07 |
| AP4M1[ad] | 7:100101549-100110345 | 4.87 | 1.12e-06 | 4.87 | 1.10e-06 | - | - | - | - |
| GAL3ST4[bd] | 7:100159244-100168617 | 5.33 | 1.01e-07 | -5.07 | 3.97e-07 | 5.29 | 1.20e-07 | 5.11 | 3.25e-07 |
| PVRIG[bd] | 7:100218241-100221490 | 5.31 | 1.08e-07 | 5.32 | 1.06e-07 | - | - | - | - |
| AC092849.1[bd] | 7:100435257-100436510 | 6.54 | 6.23e-11 | - | - | 6.48 | 9.02e-11 | 5.98 | 2.21e-09 |
| PPP1R35[bd] | 7:100435282-100436497 | 5.86 | 4.67e-09 | 4.93 | 8.40e-07 | - | - | 5.74 | 9.36e-09 |
| UFSP1[bd] | 7:100888721-100889715 | -5.67 | 1.41e-08 | -5.68 | 1.34e-08 | - | - | - | - |

| | | Avg-valid+SR ROSMAP-SMA | | SR-TWAS ROSMAP-SMA | | PrediXcan ROSMAP-SMA | | TIGAR ROSMAP-SMA | |
|---|---|---|---|---|---|---|---|---|---|
| Gene | Locus | Zscore | Pvalue | Zscore | Pvalue | Zscore | Pvalue | Zscore | Pvalue |
| FAM13C[bd] | 10:59246130-59363181 | -4.85 | 1.21e-06 | -4.87 | 1.13e-06 | - | - | - | - |
| RN7SL435P[bd] | 11:59291052-59291333 | 7.33 | 2.36e-13 | - | - | - | - | 7.33 | 2.36e-13 |
| AP000640.2[bd] | 11:59669312-59676041 | 5.84 | 5.19e-09 | - | - | - | - | 5.84 | 5.19e-09 |
| SRD5A3P1[bd] | 11:59898185-59899133 | 8.73 | 2.49e-18 | - | - | - | - | 8.73 | 2.49e-18 |
| AP000790.1[bd] | 11:59942879-59970146 | -5.47 | 4.41e-08 | -5.67 | 1.41e-08 | - | - | - | - |
| LINC02705[ad] | 11:60159687-60160822 | -5.65 | 1.56e-08 | - | - | - | - | -5.65 | 1.56e-08 |
| MS4A4E[bd] | 11:60200270-60243137 | -8.59 | 8.94e-18 | - | - | - | - | -8.59 | 8.94e-18 |
| MS4A8[bd] | 11:60699585-60715807 | -5.12 | 3.14e-07 | - | - | - | - | - | - |
| MS4A10[bd] | 11:60785333-60801305 | -5.88 | 4.10e-09 | - | - | - | - | -5.88 | 4.10e-09 |
| AP003721.1[bd] | 11:60916339-60925397 | 7.72 | 1.20e-14 | - | - | - | - | 7.41 | 1.24e-13 |
| SLC15A3[bd] | 11:60937084-60952530 | -4.84 | 1.31e-06 | -7.57 | 3.82e-14 | - | - | -4.74 | 2.11e-06 |
| AP002803.1[bd] | 11:85336075-85336572 | -6.69 | 2.18e-11 | - | - | - | - | -6.69 | 2.18e-11 |
| AP003128.1[bd] | 11:85916502-85923682 | -6.83 | 8.46e-12 | - | - | - | - | -6.83 | 8.46e-12 |
| RN7SL225P[bd] | 11:86324012-86324309 | 5.05 | 4.33e-07 | - | - | 5.03 | 5.02e-07 | 4.93 | 8.04e-07 |
| LINC02833[bd] | 14:92886352-92893508 | -5.16 | 2.44e-07 | - | - | - | - | -5.16 | 2.44e-07 |
| AL110118.2[bd] | 14:93184973-93218586 | -5.32 | 1.01e-07 | - | - | -5.25 | 1.52e-07 | -5.63 | 1.77e-08 |
| RN7SL819P[bd] | 17:44718861-44719130 | -5.31 | 1.07e-07 | - | - | - | - | -5.31 | 1.07e-07 |
| PPP1R9B[bd] | 17:50133737-50150677 | 5.07 | 3.93e-07 | 5.11 | 3.15e-07 | - | - | - | - |
| ACE[ac] | 17:63477061-63498380 | 5.05 | 4.42e-07 | 5.05 | 4.44e-07 | - | - | - | - |
| KCNN4[bd] | 19:43766533-43781257 | 5.34 | 9.37e-08 | - | - | 5.42 | 6.04e-08 | - | - |
| AC006213.5[bd] | 19:43978376-43978663 | 8.33 | 8.19e-17 | 11.88 | 1.43e-32 | 7.48 | 7.22e-14 | 5.55 | 2.85e-08 |
| AC006213.1[bd] | 19:43996896-44003298 | 7.53 | 5.24e-14 | 6.47 | 1.01e-10 | - | - | 4.78 | 1.74e-06 |
| AC067968.1[bd] | 19:44025354-44087318 | 5.52 | 3.36e-08 | - | - | - | - | 5.52 | 3.36e-08 |
| ZNF227[bd] | 19:44207547-44237268 | -6.1 | 1.03e-09 | -6.14 | 8.16e-10 | - | - | - | - |
| AC138470.1[bd] | 19:44246478-44248374 | -7.05 | 1.79e-12 | -6.76 | 1.42e-11 | - | - | -9.21 | 3.26e-20 |
| ZNF112[bd] | 19:44326555-44367217 | -10.24 | 1.26e-24 | -10.25 | 1.24e-24 | - | - | - | - |
| ZNF229[bc] | 19:44417519-44448578 | 8.98 | 2.63e-19 | 8.99 | 2.58e-19 | - | - | - | - |
| AC243964.3[bd] | 19:44631573-44725217 | -10.53 | 5.98e-26 | -10.53 | 6.02e-26 | - | - | - | - |
| PVR[ac] | 19:44643798-44666162 | 6.51 | 7.45e-11 | 6.51 | 7.45e-11 | - | - | - | - |
| CEACAM19[ac] | 19:44662278-44684359 | -16.26 | 1.84e-59 | -16.3 | 9.48e-60 | - | - | - | - |

| | | Avg-valid+SR ROSMAP-SMA | | SR-TWAS ROSMAP-SMA | | PrediXcan ROSMAP-SMA | | TIGAR ROSMAP-SMA | |
|---|---|---|---|---|---|---|---|---|---|
| Gene | Locus | Zscore | Pvalue | Zscore | Pvalue | Zscore | Pvalue | Zscore | Pvalue |
| AC243964.4[bd] | 19:44664131-44666158 | 7.02 | 2.16e-12 | - | - | - | - | 7.02 | 2.16e-12 |
| TOMM40[ac] | 19:44890569-44903689 | -9.21 | 3.31e-20 | - | - | - | - | -9.21 | 3.31e-20 |
| AC011481.3[bd] | 19:44909375-44914968 | -4.88 | 1.04e-06 | - | - | - | - | -4.88 | 1.04e-06 |
| APOC1[ac] | 19:44914247-44919349 | -6.68 | 2.36e-11 | -6.54 | 6.12e-11 | - | - | -16.86 | 9.08e-64 |
| APOC2[ac] | 19:44945971-44949566 | -10.58 | 3.52e-26 | -10.99 | 4.23e-28 | - | - | 14.64 | 1.53e-48 |
| AC011481.1[bd] | 19:44950044-44954007 | 5.66 | 1.53e-08 | - | - | - | - | 9.13 | 7.03e-20 |
| CLASRP[ad] | 19:45039045-45070956 | -9.96 | 2.23e-23 | -13.52 | 1.21e-41 | - | - | - | - |
| GEMIN7-AS1[ac] | 19:45076510-45092635 | -11.85 | 2.21e-32 | - | - | -12 | 3.51e-33 | - | - |
| GEMIN7[ac] | 19:45079195-45091518 | -7.4 | 1.36e-13 | -7.4 | 1.38e-13 | - | - | -8.38 | 5.41e-17 |
| MARK4[ad] | 19:45079288-45305284 | -5.84 | 5.16e-09 | -6.53 | 6.76e-11 | - | - | -5.77 | 7.88e-09 |
| PPP1R37[ad] | 19:45091396-45148077 | 4.83 | 1.35e-06 | - | - | - | - | 9.19 | 3.81e-20 |
| TRAPPC6A[ac] | 19:45162928-45178237 | 16.77 | 3.89e-63 | 16.39 | 2.42e-60 | - | - | - | - |
| BLOC1S3[ac] | 19:45178784-45216933 | -25.25 | 1.22e-140 | -4.94 | 7.79e-07 | -25.92 | 3.97e-148 | -12.14 | 6.46e-34 |
| EXOC3L2[ad] | 19:45212621-45245431 | 9.56 | 1.20e-21 | - | - | - | - | 9.56 | 1.20e-21 |
| FOSB[bd] | 19:45467995-45475179 | 29.19 | 2.44e-187 | - | - | 29.2 | 1.87e-187 | 13.79 | 2.98e-43 |
| RTN2[bd] | 19:45485294-45497055 | -5.5 | 3.89e-08 | - | - | - | - | -5.29 | 1.23e-07 |
| GPR4[bd] | 19:45589764-45602212 | -30.87 | 3.48e-209 | - | - | -30.86 | 4.04e-209 | -8.43 | 3.53e-17 |
| QPCTL[bd] | 19:45692666-45703989 | -4.94 | 7.79e-07 | -4.92 | 8.79e-07 | - | - | -7.2 | 6.20e-13 |
| FBXO46[bd] | 19:45710629-45730896 | 5.33 | 9.84e-08 | - | - | 5.41 | 6.37e-08 | - | - |
| AC007191.1[bd] | 19:45714387-45717381 | -16.79 | 2.85e-63 | - | - | - | - | -16.79 | 2.85e-63 |
| DM1-AS[bd] | 19:45767796-45772504 | 10.06 | 8.33e-24 | 6.66 | 2.73e-11 | - | - | 9.92 | 3.57e-23 |
| DMPK[bd] | 19:45769717-45782552 | -4.88 | 1.06e-06 | 16.52 | 2.79e-61 | -4.79 | 1.65e-06 | -11.18 | 5.21e-29 |
| AC011530.1[bd] | 19:45779437-45785973 | -4.74 | 2.10e-06 | - | - | -4.74 | 2.18e-06 | -6.38 | 1.72e-10 |
| DMWD[bd] | 19:45782947-45792845 | 5.63 | 1.76e-08 | - | - | - | - | 5.56 | 2.67e-08 |
| AC092301.1[bd] | 19:45830164-45831108 | -9.74 | 2.07e-22 | - | - | -9.65 | 5.10e-22 | -12.03 | 2.62e-33 |
| MYPOP[bd] | 19:45890023-45902613 | -5.67 | 1.42e-08 | - | - | - | - | -8.16 | 3.43e-16 |
| AC006262.2[bd] | 19:46163893-46180647 | 5.88 | 4.02e-09 | - | - | - | - | 5.88 | 4.02e-09 |

| | | Avg-valid+SR ROSMAP-SMA | | SR-TWAS ROSMAP-SMA | | PrediXcan ROSMAP-SMA | | TIGAR ROSMAP-SMA | |
|---|---|---|---|---|---|---|---|---|---|
| Gene | Locus | Zscore | Pvalue | Zscore | Pvalue | Zscore | Pvalue | Zscore | Pvalue |

*a*: Known GWAS risk gene of AD.

*b*: Gene within 1Mb of known GWAS risk gene of AD.

*c*: Previously observed TWAS risk gene of AD.

*d*: Gene within 1MB of previously observed TWAS risk gene of AD.

**Supplementary Table 5: Significant TWAS risk genes of Parkinson's disease identified by Avg-valid+SR models for the GTEx validation data of the BRNSNG tissue. TWAS Zscores and p-values were presented here for the models of Avg-valid+SR, SR-TWAS, PrediXcan base validation models, and TIGAR base validation models. The signs of Zscores show the directions of the mediated genetic effects on AD dementia.**

| | | Avg-valid+SR GTEx-BRNSNG | | SR-TWAS GTEx-BRNSNG | | PrediXcan GTEx-BRNSNG | | TIGAR GTEx-BRNSNG | |
|---|---|---|---|---|---|---|---|---|---|
| Gene | Locus | Zscore | Pvalue | Zscore | Pvalue | Zscore | Pvalue | Zscore | Pvalue |
| PARL[bd] | 3:183829271-183884923 | -4.75 | 2.02e-06 | - | - | - | - | - | - |
| IDUA[bd] | 4:986997-1004506 | 5.58 | 2.35e-08 | 5.77 | 7.98e-09 | - | - | - | - |
| CD38[ac] | 4:15778275-15853230 | 7.59 | 3.31e-14 | 7.68 | 1.57e-14 | - | - | - | - |
| MMRN1[ac] | 4:89879532-89954629 | -7.99 | 1.34e-15 | -7.91 | 2.55e-15 | -7.9 | 2.80e-15 | -8.5 | 1.95e-17 |
| NDUFAF2[ac] | 5:60945129-61153037 | -5.33 | 9.80e-08 | -5.45 | 4.91e-08 | - | - | - | - |
| KLHL7-AS1[bc] | 7:23101228-23105703 | 5.11 | 3.19e-07 | 4.88 | 1.04e-06 | 5.11 | 3.24e-07 | 5.06 | 4.26e-07 |
| NUPL2[bc] | 7:23181827-23201009 | -5.78 | 7.51e-09 | -4.94 | 7.90e-07 | -5.8 | 6.56e-09 | - | - |
| AC005082.12[bd] | 7:23206013-23208045 | 5.94 | 2.83e-09 | 5.47 | 4.44e-08 | - | - | 5.35 | 8.66e-08 |
| GPNMB[ac] | 7:23235967-23275108 | -5.47 | 4.39e-08 | -5.52 | 3.31e-08 | - | - | -5.11 | 3.28e-07 |
| LA16c-431H6.7 | 16:1688355-1690536 | -4.95 | 7.60e-07 | - | - | - | - | - | - |
| MYLPF[bd] | 16:30370934-30377991 | 6.34 | 2.31e-10 | - | - | - | - | 6.31 | 2.78e-10 |
| ZNF688[bd] | 16:30569346-30572734 | -5.16 | 2.42e-07 | -5.14 | 2.71e-07 | - | - | - | - |
| PRR14[bd] | 16:30650729-30656440 | -4.97 | 6.71e-07 | -4.92 | 8.80e-07 | - | - | - | - |
| CCDC189[bd] | 16:30761198-30762710 | -4.79 | 1.64e-06 | - | - | - | - | - | - |
| RP11-2C24.7[bd] | 16:30821513-30821884 | 5.19 | 2.14e-07 | 5.22 | 1.81e-07 | - | - | - | - |
| RP11-1072A3.3[bd] | 16:30984665-30987767 | -5.13 | 2.92e-07 | -4.92 | 8.63e-07 | - | - | -4.99 | 5.90e-07 |
| RP11-196G11.6[bd] | 16:31056460-31062803 | -5.74 | 9.39e-09 | -5.55 | 2.90e-08 | - | - | -5.33 | 9.79e-08 |
| ZNF668[bc] | 16:31063813-31074320 | -5.27 | 1.36e-07 | -5.37 | 8.01e-08 | - | - | - | - |
| PRSS53[ac] | 16:31084197-31088963 | -4.93 | 8.42e-07 | -5.05 | 4.41e-07 | - | - | -4.88 | 1.04e-06 |
| VKORC1[bc] | 16:31090842-31095980 | -4.8 | 1.60e-06 | -5.56 | 2.75e-08 | - | - | - | - |
| KAT8[bd] | 16:31115754-31131393 | 5.06 | 4.18e-07 | 5.16 | 2.42e-07 | - | - | - | - |
| PRSS36[bd] | 16:31138925-31150094 | 4.84 | 1.29e-06 | - | - | - | - | - | - |
| AHSP[bd] | 16:31527864-31528803 | 5.18 | 2.20e-07 | 5.17 | 2.31e-07 | - | - | 4.77 | 1.80e-06 |
| RP11-1166P10.1[bd] | 16:31975803-32003728 | -4.81 | 1.53e-06 | -4.8 | 1.63e-06 | - | - | - | - |
| ADORA2B[d] | 17:15944917-15975746 | -5.02 | 5.16e-07 | -5.2 | 2.04e-07 | -4.94 | 7.96e-07 | -5.29 | 1.21e-07 |

| | | Avg-valid+SR GTEx-BRNSNG | | SR-TWAS GTEx-BRNSNG | | PrediXcan GTEx-BRNSNG | | TIGAR GTEx-BRNSNG | |
|---|---|---|---|---|---|---|---|---|---|
| Gene | Locus | Zscore | Pvalue | Zscore | Pvalue | Zscore | Pvalue | Zscore | Pvalue |
| HIGD1B[bd] | 17:44846353-44849388 | -5.68 | 1.35e-08 | -5.71 | 1.16e-08 | - | - | - | - |
| EFTUD2[bd] | 17:44849943-44899662 | -5.87 | 4.44e-09 | -7.15 | 8.77e-13 | - | - | -5.83 | 5.43e-09 |
| KIF18B[bd] | 17:44924709-44947711 | -7.06 | 1.71e-12 | - | - | - | - | -7.42 | 1.15e-13 |
| PLCD3[bd] | 17:45109017-45133354 | -5.58 | 2.39e-08 | -5.72 | 1.08e-08 | - | - | 5.62 | 1.87e-08 |
| ACBD4[bd] | 17:45132600-45144181 | 7.02 | 2.25e-12 | - | - | - | - | 7.31 | 2.63e-13 |
| AC142472.6[bd] | 17:45146730-45148470 | -5.14 | 2.75e-07 | - | - | - | - | -5.07 | 3.94e-07 |
| HEXIM1[bd] | 17:45148502-45152101 | -7.5 | 6.56e-14 | - | - | - | - | -8.27 | 1.34e-16 |
| HEXIM2[bd] | 17:45160700-45162859 | 8.94 | 3.88e-19 | -7.75 | 9.24e-15 | - | - | 8.93 | 4.20e-19 |
| CTD-2020K17.1[bd] | 17:45190931-45221788 | 8.5 | 1.92e-17 | 7.45 | 9.61e-14 | - | - | 8.49 | 2.14e-17 |
| FMNL1[bd] | 17:45222223-45247266 | 7.67 | 1.70e-14 | 7.53 | 5.26e-14 | - | - | 6.85 | 7.20e-12 |
| MAP3K14-AS1[bd] | 17:45249534-45268630 | 5.01 | 5.50e-07 | 5.24 | 1.61e-07 | - | - | -5.7 | 1.16e-08 |
| MAP3K14[bd] | 17:45263392-45317040 | 7.6 | 3.01e-14 | 5.18 | 2.27e-07 | - | - | 8 | 1.24e-15 |
| ARHGAP27[ad] | 17:45393902-45434421 | 8.67 | 4.20e-18 | 6.59 | 4.49e-11 | - | - | 8.73 | 2.54e-18 |
| CTB-39G8.3[bd] | 17:45397025-45397477 | -8.63 | 6.02e-18 | - | - | - | - | -8.87 | 7.35e-19 |
| PLEKHM1[ad] | 17:45435900-45490749 | 7.73 | 1.06e-14 | 7.22 | 5.12e-13 | - | - | 8.41 | 4.21e-17 |
| LRRC37A4P[bd] | 17:45506741-45550335 | -9.28 | 1.66e-20 | -9.41 | 4.98e-21 | - | - | -9.13 | 6.85e-20 |
| DND1P1[bd] | 17:45585871-45586929 | 9.27 | 1.86e-20 | 9.14 | 6.32e-20 | - | - | 9.32 | 1.15e-20 |
| RP11-707O23.1[bd] | 17:45592621-45593369 | 9.04 | 1.55e-19 | 9.04 | 1.54e-19 | - | - | - | - |
| MAPK8IP1P2[bd] | 17:45600869-45602340 | 9.31 | 1.30e-20 | 9.05 | 1.39e-19 | - | - | 9.42 | 4.38e-21 |
| LINC02210[ad] | 17:45620328-45655156 | 9.2 | 3.44e-20 | 9.06 | 1.30e-19 | - | - | 9.2 | 3.44e-20 |
| MAPT-AS1[ad] | 17:45799390-45895630 | -7.07 | 1.53e-12 | -6.65 | 2.97e-11 | - | - | -7.05 | 1.75e-12 |
| MAPT[ac] | 17:45894382-46028334 | 9.14 | 6.29e-20 | 9.14 | 6.26e-20 | - | - | - | - |
| MAPT-IT1[bd] | 17:45895783-45898798 | 8.11 | 5.03e-16 | 7.62 | 2.48e-14 | - | - | 8.11 | 5.00e-16 |
| STH[bd] | 17:45999250-45999694 | 7.89 | 3.07e-15 | 6.89 | 5.70e-12 | - | - | 7.88 | 3.40e-15 |
| RP11-669E14.6[bd] | 17:46035521-46035770 | 8.04 | 9.06e-16 | - | - | - | - | 7.2 | 6.12e-13 |
| KANSL1-AS1[bd] | 17:46193576-46196723 | 8.64 | 5.51e-18 | 8.12 | 4.75e-16 | - | - | 8.84 | 9.22e-19 |
| MAPK8IP1P1[bd] | 17:46243606-46245044 | 9.06 | 1.27e-19 | 9.15 | 5.49e-20 | - | - | 8.8 | 1.34e-18 |
| RP11-259G18.3[bd] | 17:46259551-46260606 | 9.27 | 1.87e-20 | 8.98 | 2.60e-19 | - | - | 9.39 | 5.90e-21 |
| RP11-259G18.1[bd] | 17:46267037-46268694 | 9.14 | 6.29e-20 | 9.11 | 8.37e-20 | - | - | 9.06 | 1.36e-19 |
| LRRC37A2[bc] | 17:46511511-46553449 | 8.05 | 8.31e-16 | 6.77 | 1.33e-11 | 8.01 | 1.18e-15 | 9.19 | 3.85e-20 |

| | | Avg-valid+SR GTEx-BRNSNG | | SR-TWAS GTEx-BRNSNG | | PrediXcan GTEx-BRNSNG | | TIGAR GTEx-BRNSNG | |
|---|---|---|---|---|---|---|---|---|---|
| Gene | Locus | Zscore | Pvalue | Zscore | Pvalue | Zscore | Pvalue | Zscore | Pvalue |
| ARL17A[bd] | 17:46516702-46579682 | 7.22 | 5.30e-13 | 6.27 | 3.68e-10 | - | - | 7.67 | 1.69e-14 |
| NSF[ad] | 17:46590669-46757464 | -5.34 | 9.19e-08 | -5.05 | 4.49e-07 | - | - | -8.22 | 1.97e-16 |
| RPS7P11[bd] | 17:46721582-46722167 | -6.74 | 1.56e-11 | - | - | - | - | -7.29 | 3.02e-13 |
| WNT9B[bd] | 17:46833201-46886730 | -9.06 | 1.34e-19 | -6.04 | 1.57e-09 | - | - | -8.98 | 2.71e-19 |
| RPRML[bd] | 17:46978156-46979248 | -8.68 | 4.01e-18 | 7.27 | 3.66e-13 | - | - | -8.76 | 1.88e-18 |

*a*: Known GWAS risk gene of PD.

*b*: Gene within 1MB of known GWAS risk gene of PD.

*c*: Previously observed TWAS risk gene of PD.

*d*: Gene within 1MB of previously observed TWAS risk gene of PD.

### Supplemental Figures

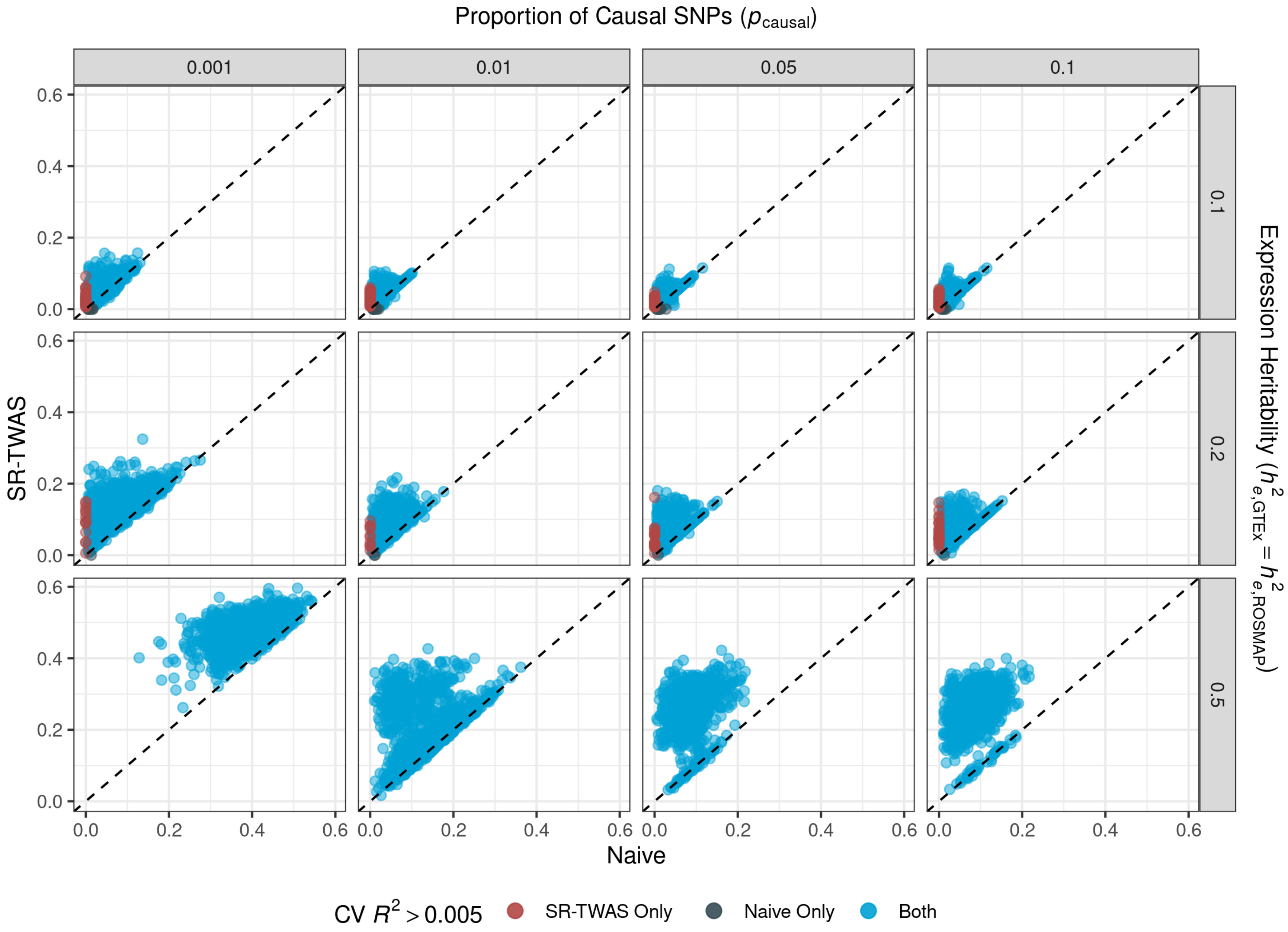

**Supplementary Figure 1: Comparison of 5-Fold CV $R^2$ by Naive and SR-TWAS methods for the scenario with GTEx causal SNPs chosen as 50% overlapped with ROSMAP causal SNPs.**

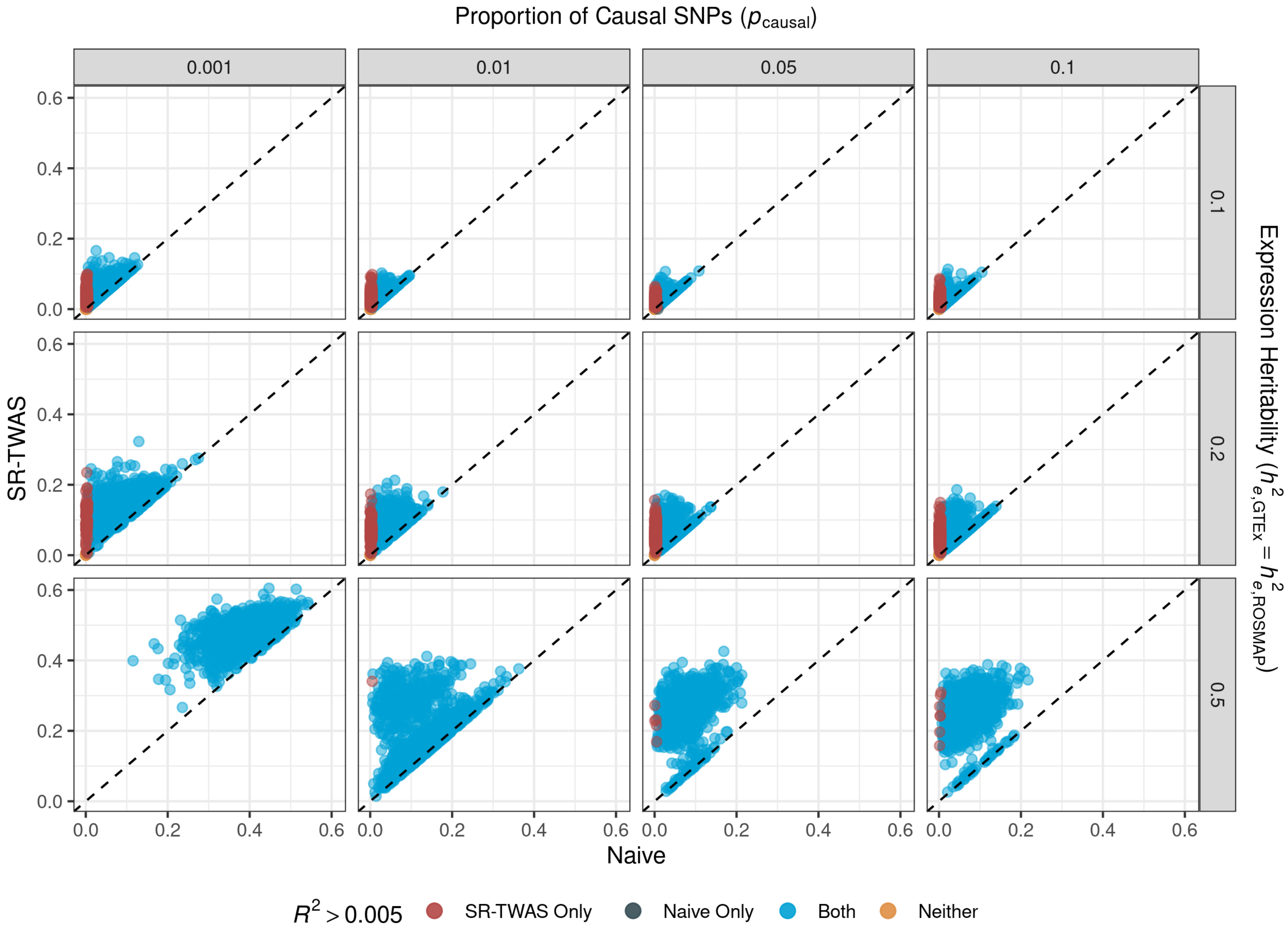

**Supplementary Figure 2: Comparison of Training $R^2$ by Naive and SR-TWAS methods for the scenario with GTEx causal SNPs chosen as 50% overlapped with ROSMAP causal SNPs.**

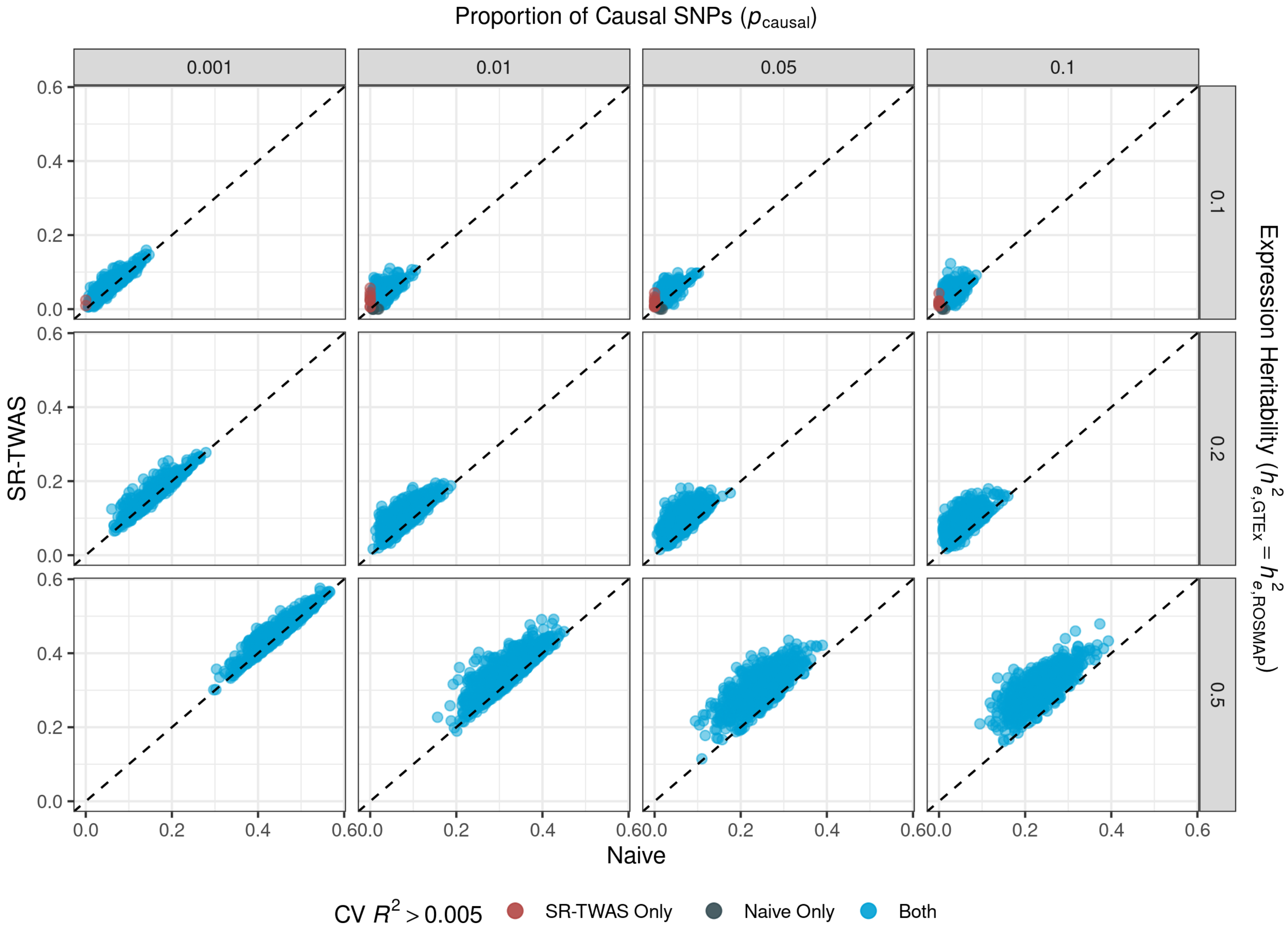

**Supplementary Figure 3: Comparison of 5-Fold CV $R^2$ by Naive and SR-TWAS methods for the scenario with GTEx expression heritability equal to that of ROSMAP ($h_{e,\mathbf{GTEx}} = h_{e,\mathbf{ROSMAP}}$), with the same set of randomly selected causal SNPs.**

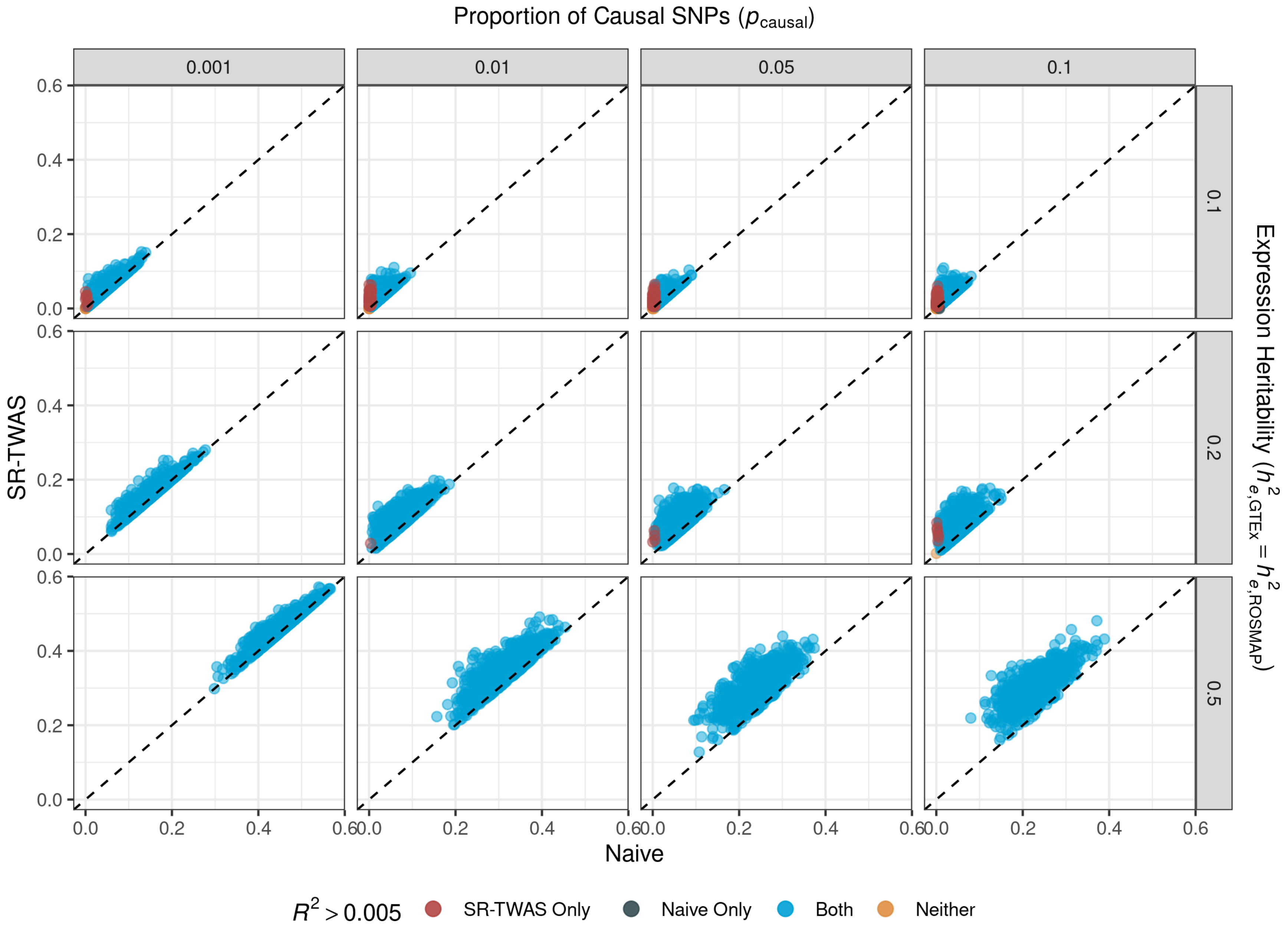

**Supplementary Figure 4: Comparison of Training $R^2$ by Naive and SR-TWAS methods for the scenario with GTEx expression heritability equal to that of ROSMAP ($h_{e,\mathbf{GTEx}} = h_{e,\mathbf{ROSMAP}}$), with the same set of randomly selected causal SNPs.**

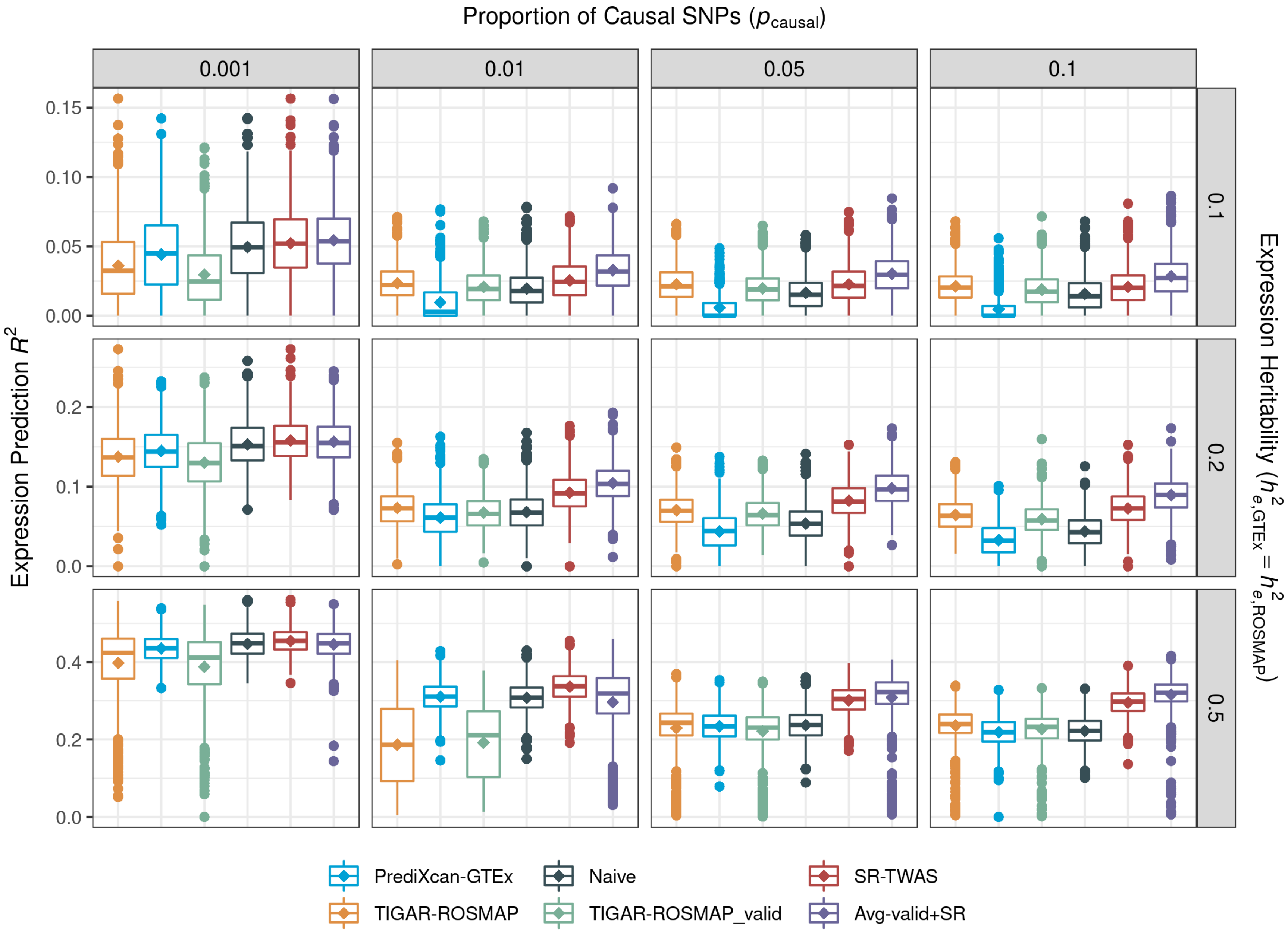

**Supplementary Figure 5: Comparison of Expression Prediction $R^2$ for the scenario with GTEx expression heritability equal to that of ROSMAP ($h_{e,\mathbf{GTEx}} = h_{e,\mathbf{ROSMAP}}$), with the same set of randomly selected causal SNPs.**

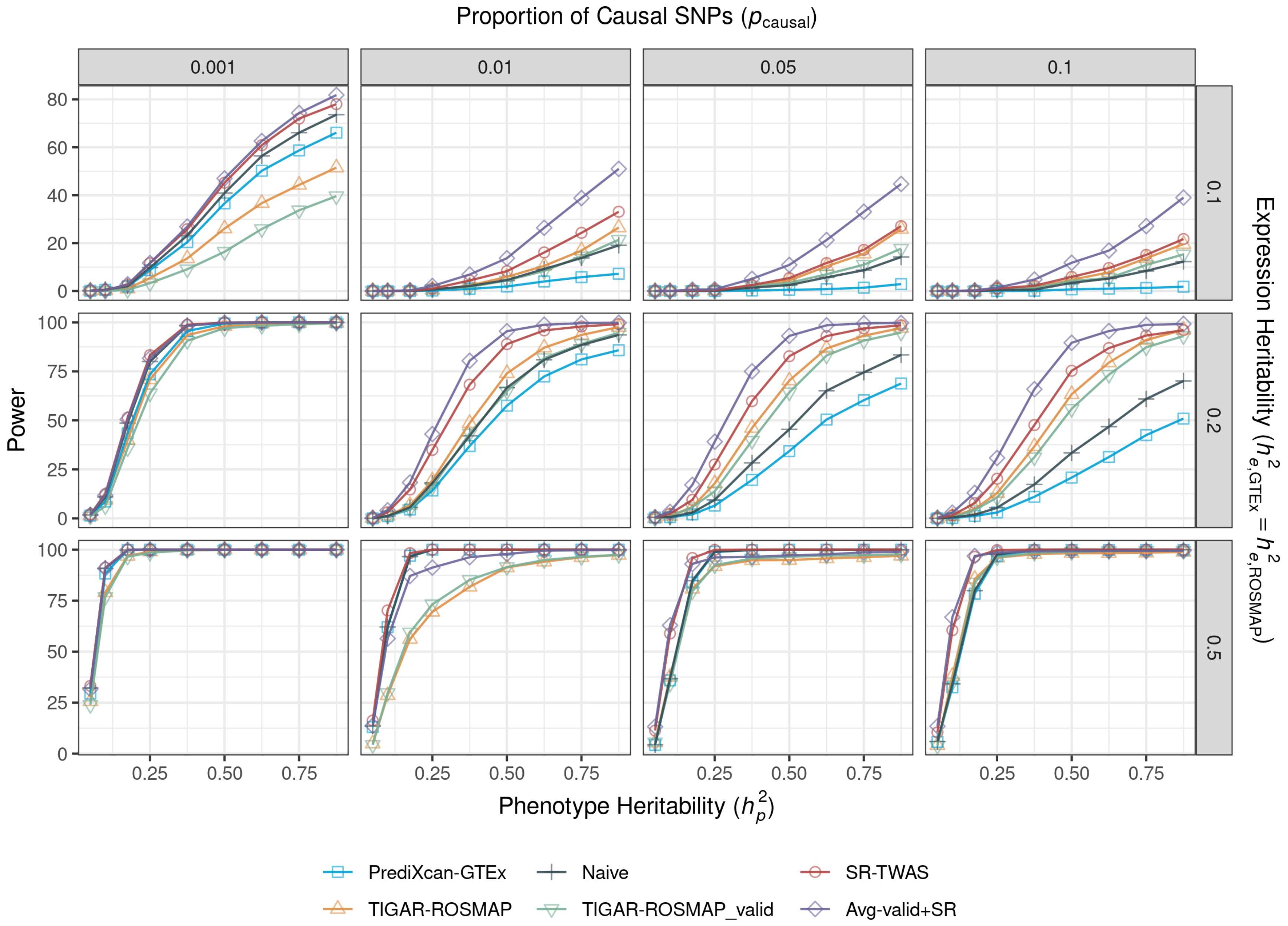

**Supplementary Figure 6: Power comparison for the scenario with GTEx expression heritability equal to that of ROSMAP ($h_{e,\mathbf{GTEx}} = h_{e,\mathbf{ROSMAP}}$), with the same set of randomly selected causal SNPs.**

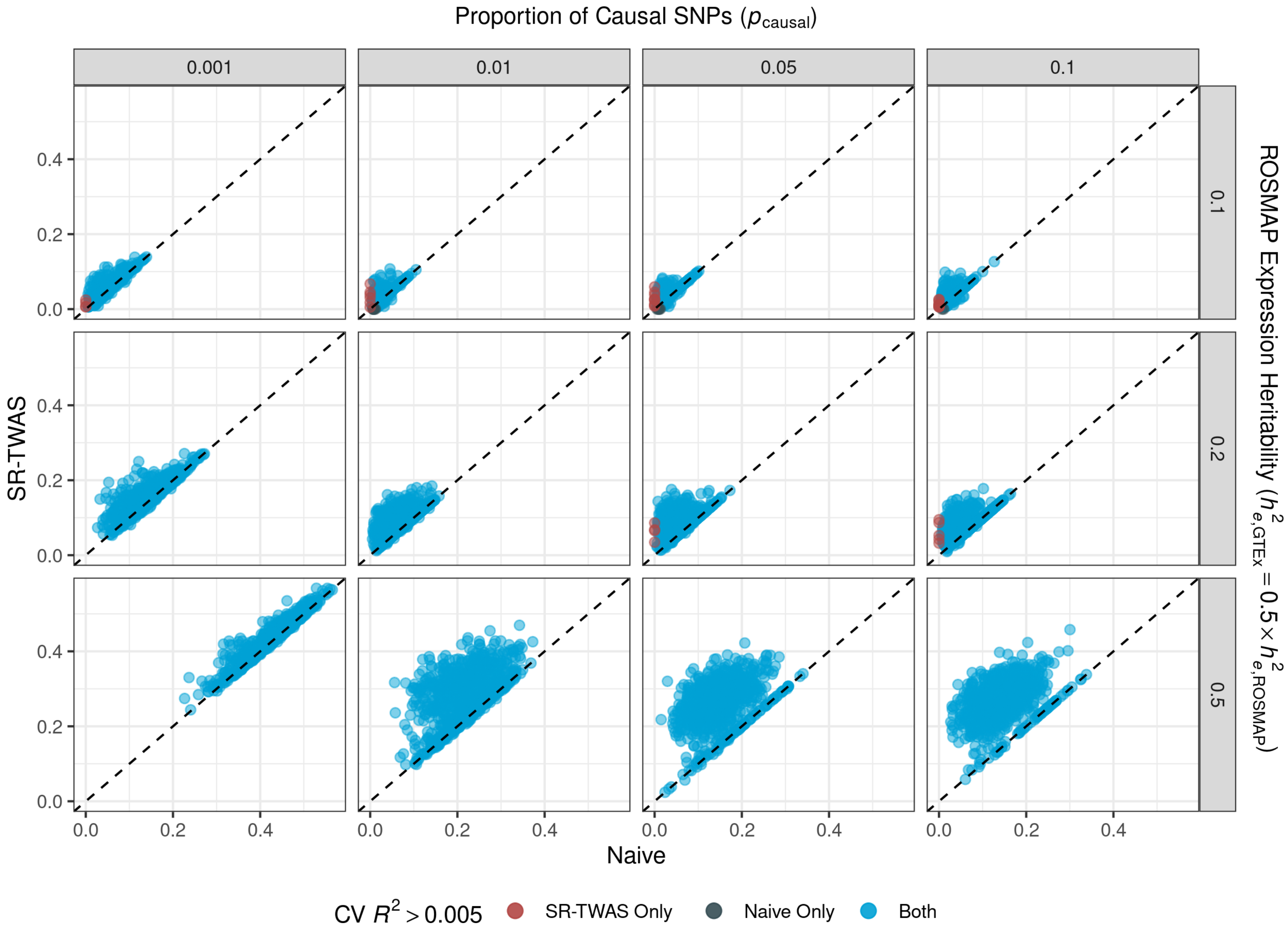

**Supplementary Figure 7: Comparison of 5-Fold CV $R^2$ by Naive and SR-TWAS methods for the scenario with GTEx expression heritability set to half that of ROSMAP ($h_{e,\mathbf{GTEx}} = 0.5 \times h_{e,\mathbf{ROSMAP}}$), with the same set of randomly selected causal SNPs.**

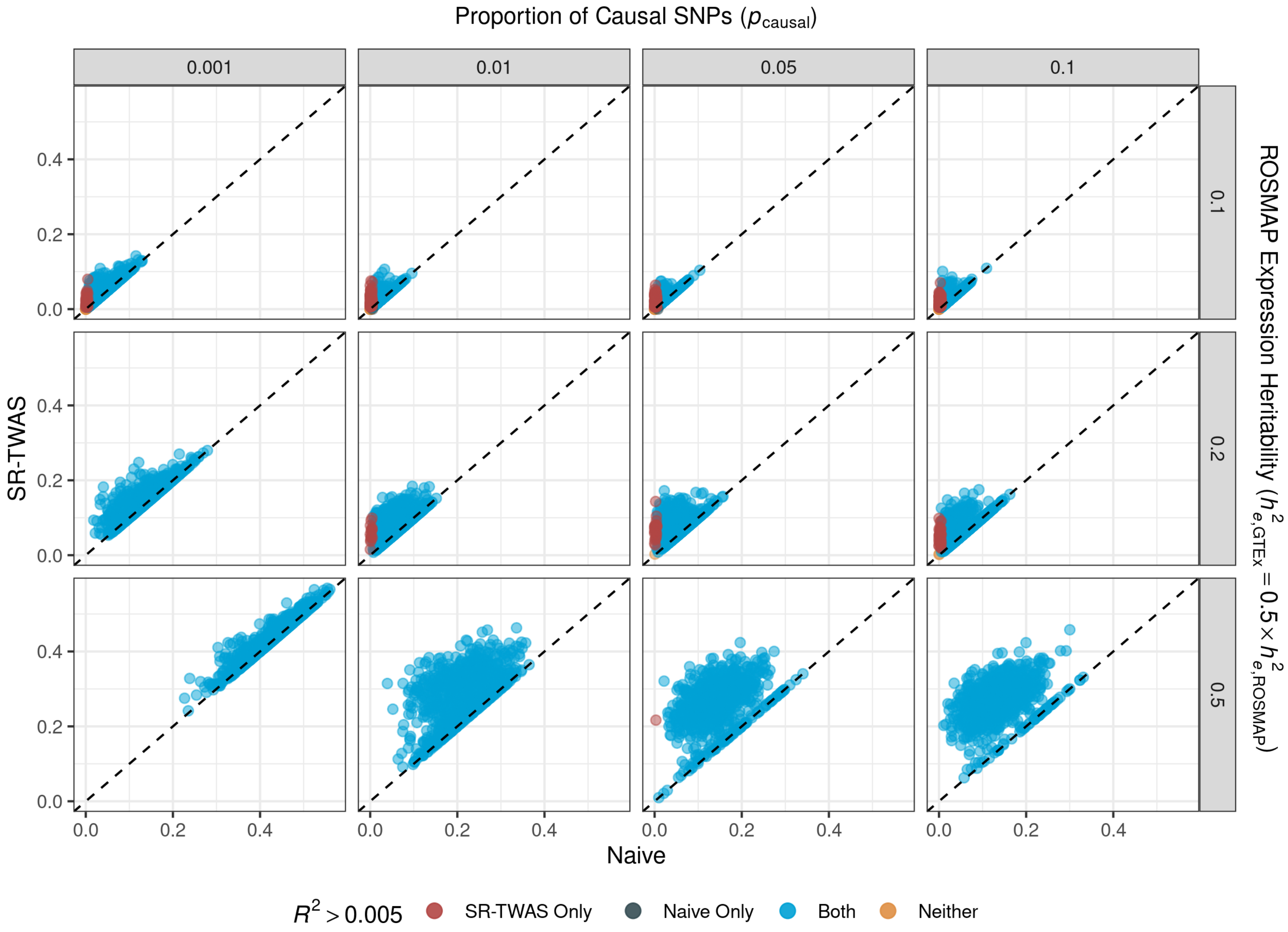

Supplementary Figure 8: Comparison of Training $R^2$ by Naive and SR-TWAS methods for the scenario with GTEx expression heritability set to half that of ROSMAP ($h_{e,\mathbf{GTEx}} = 0.5 \times h_{e,\mathbf{ROSMAP}}$), with the same set of randomly selected causal SNPs.

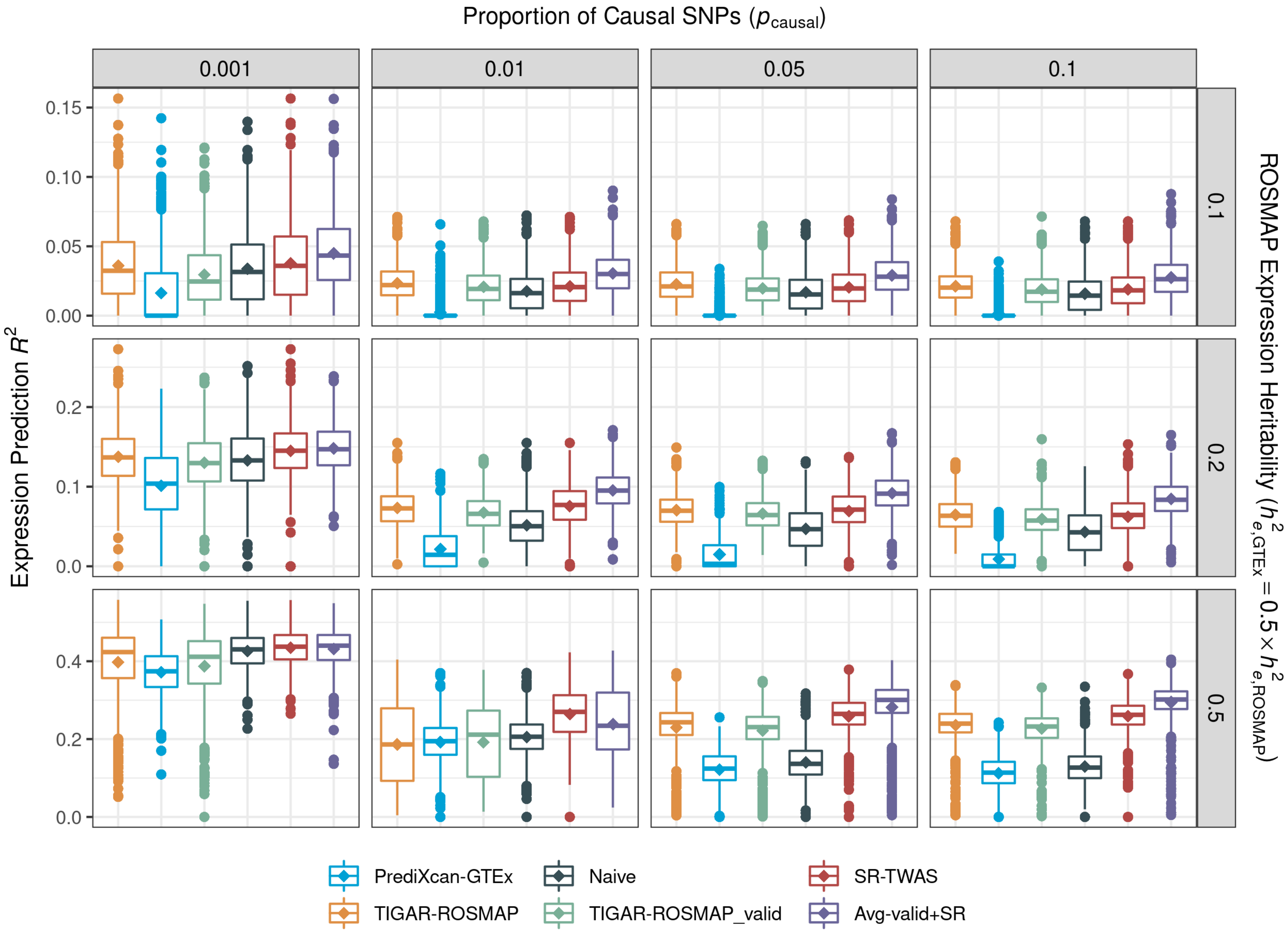

**Supplementary Figure 9: Comparison of Expression Prediction $R^2$ for the scenario with GTEx expression heritability set to half that of ROSMAP ($h_{e,\mathbf{GTEx}} = 0.5 \times h_{e,\mathbf{ROSMAP}}$), with the same set of randomly selected causal SNPs.**

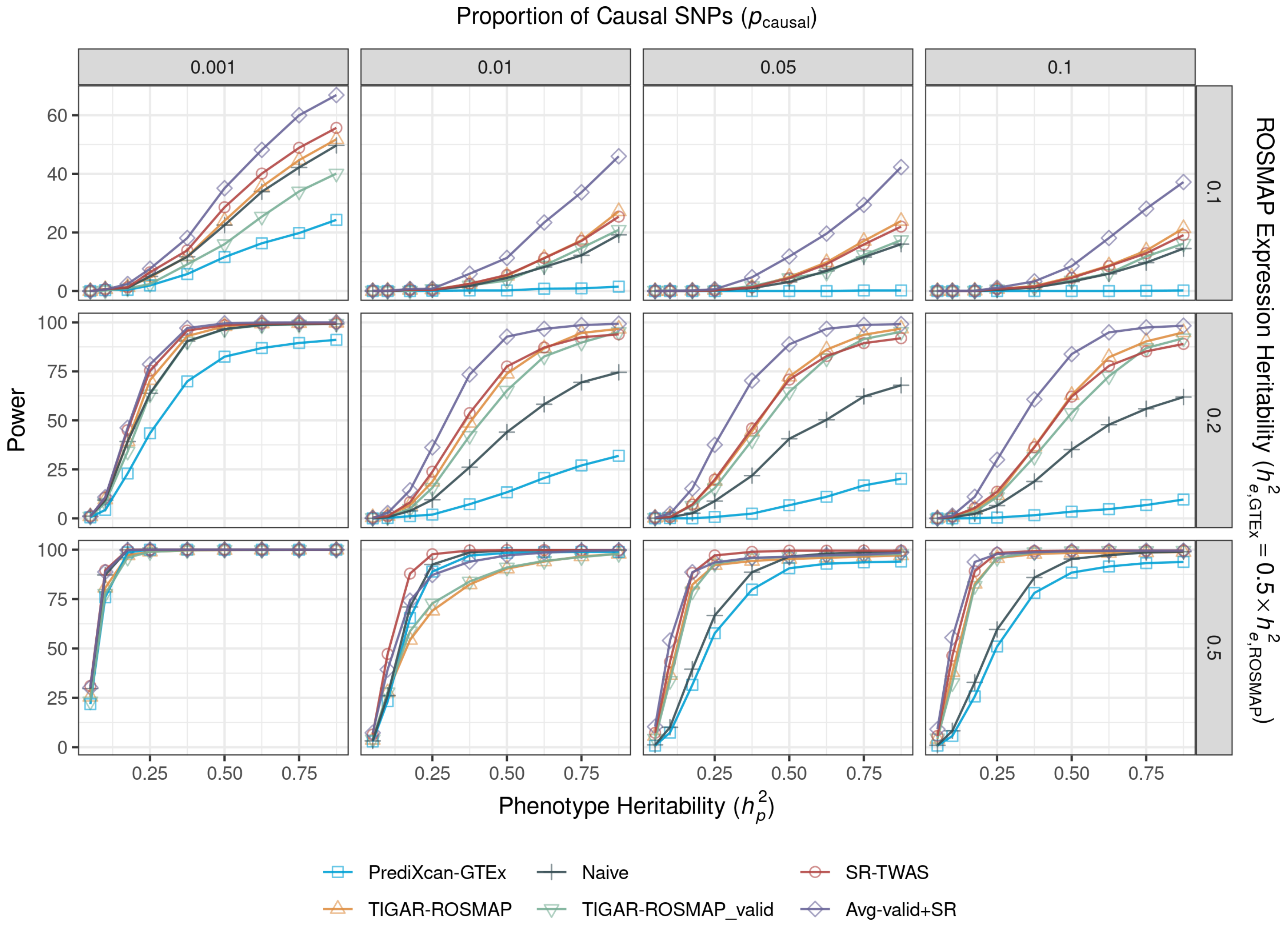

**Supplementary Figure 10: Power comparison for the scenario with GTEx expression heritability set to half that of ROSMAP ($h_{e,\mathbf{GTEx}} = 0.5 \times h_{e,\mathbf{ROSMAP}}$), with the same set of randomly selected causal SNPs.**

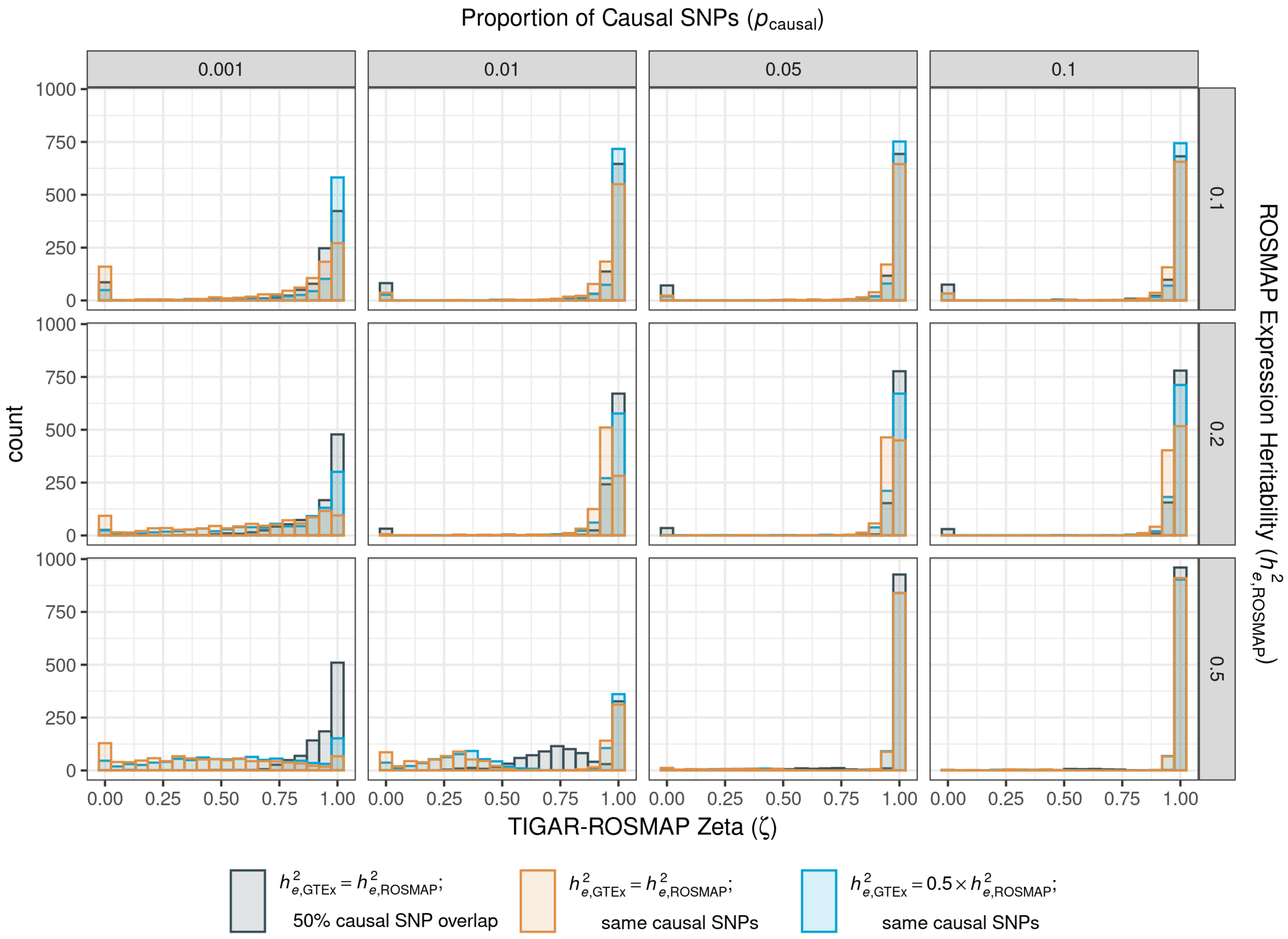

**Supplementary Figure 11: Plots of Zeta weights for base training models estimated by SR-TWAS across all simulation scenarios.**

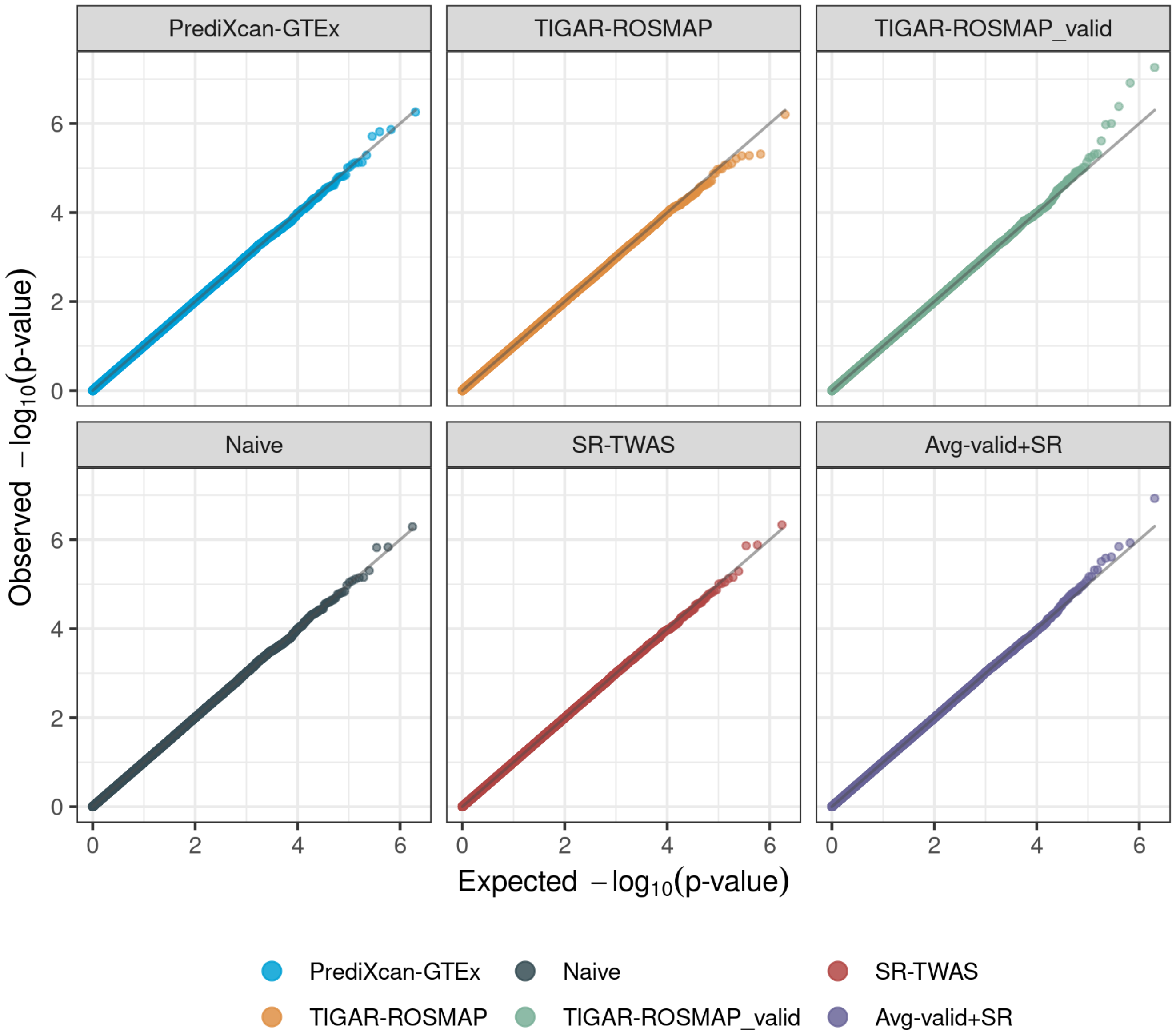

**Supplementary Figure 12: Quantile-Quantile (QQ) plots with TWAS p-values in null simulations.**

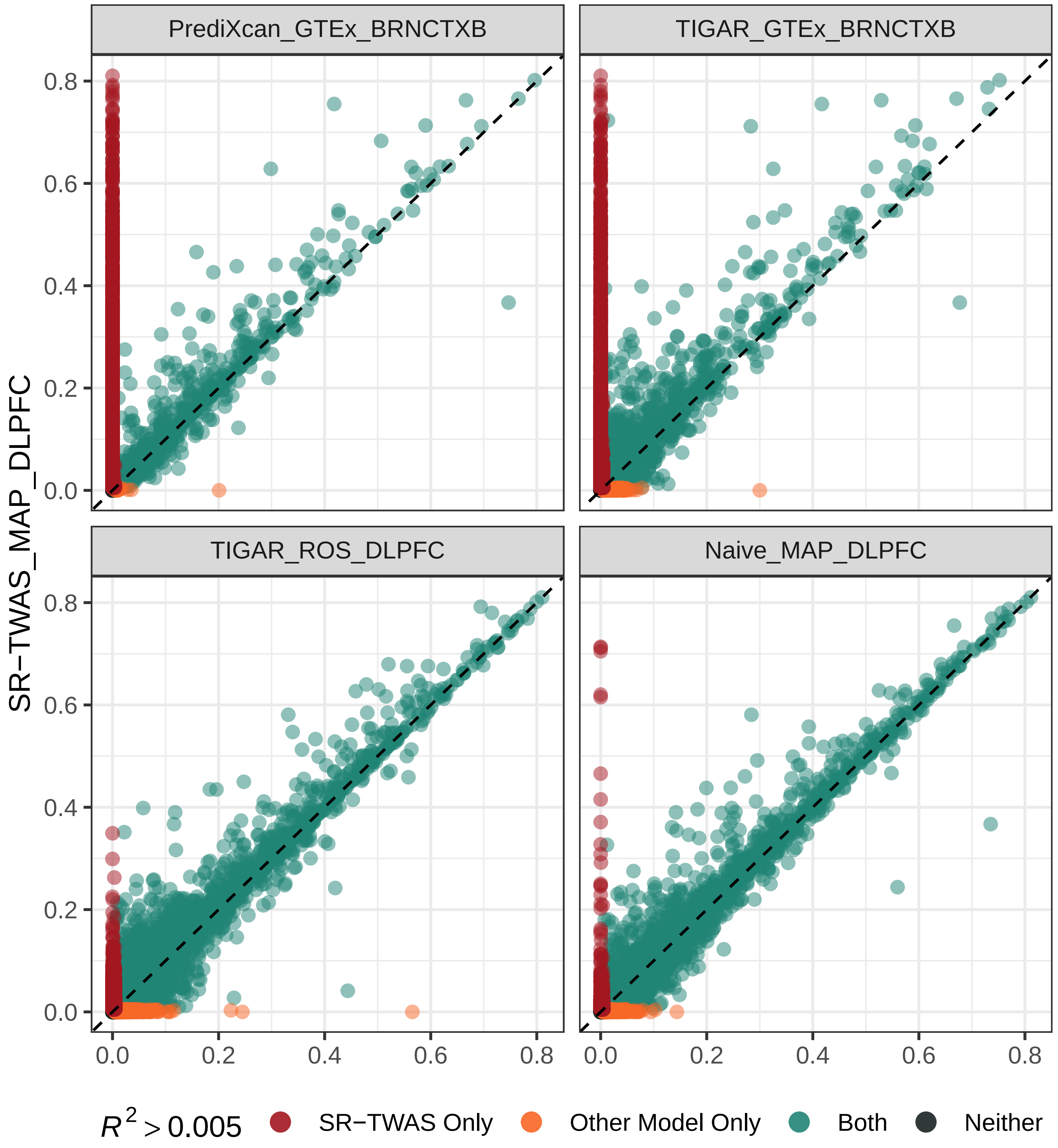

**Supplementary Figure 13: Comparison of Test $R^2$ for SR-TWAS (y-axis) and base models and Naive method (x-axis) in the real validation studies.**

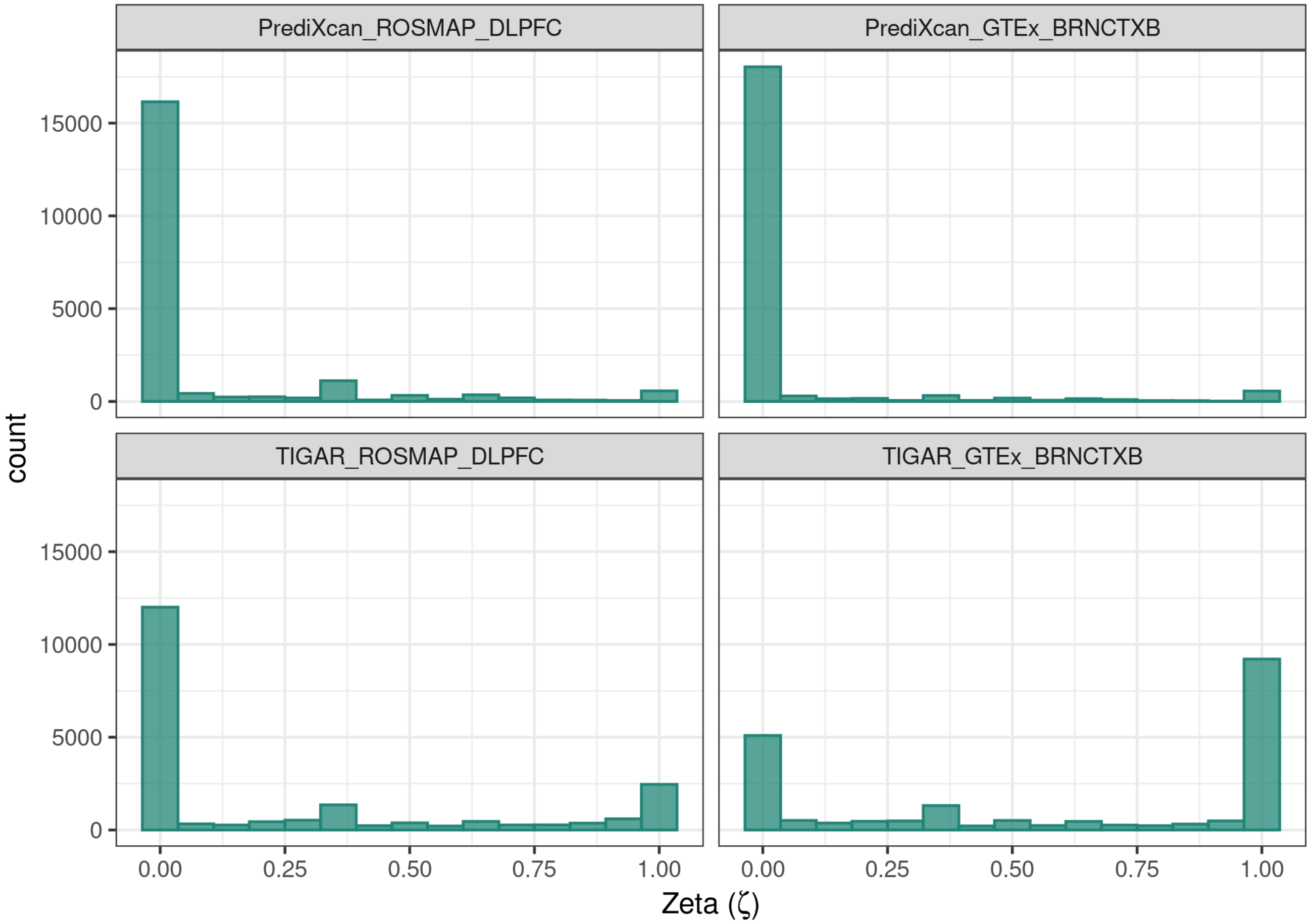

**Supplementary Figure 14: Plots of Zeta weights estimated by SR-TWAS for base training models used in AD TWAS.**

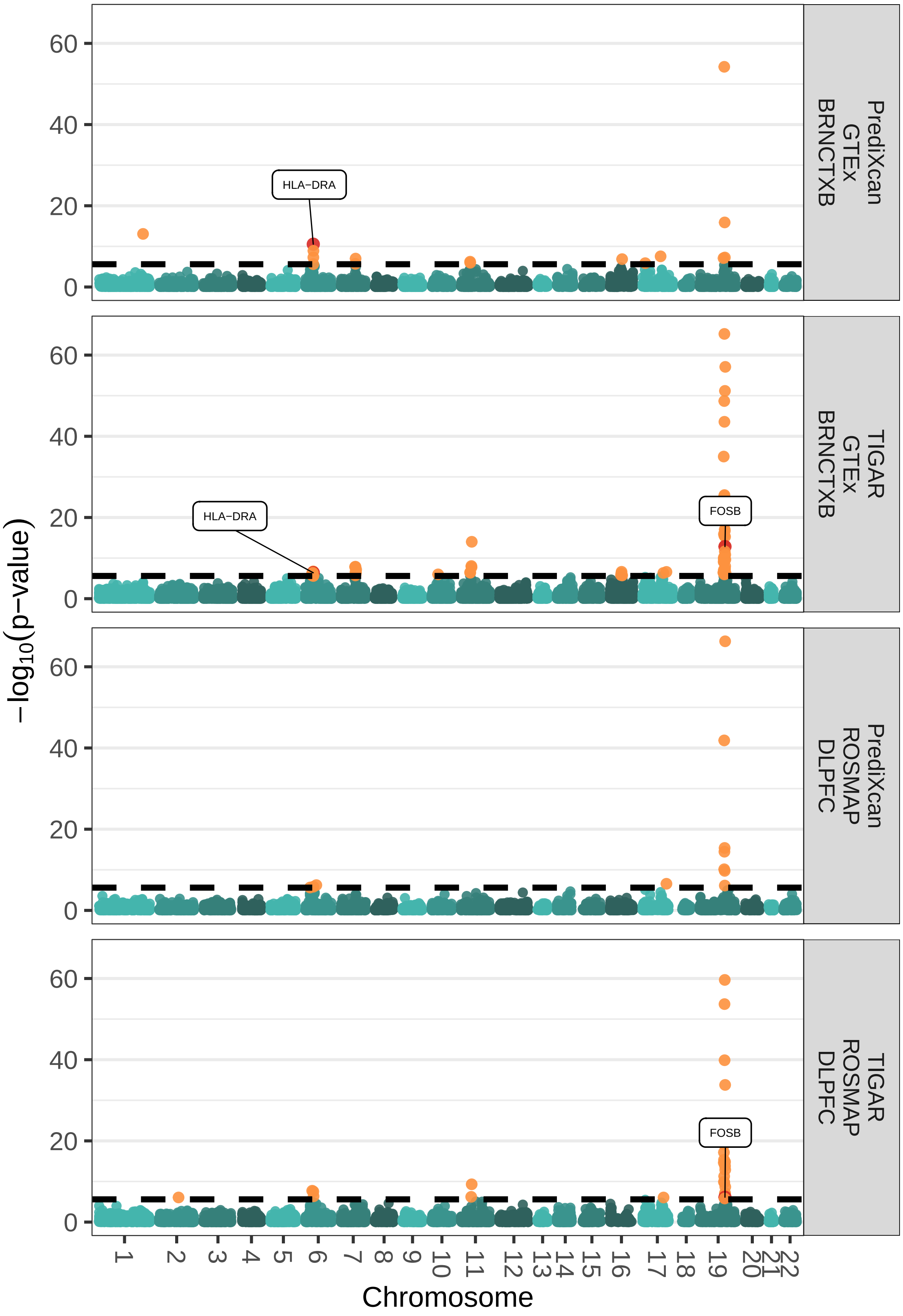

**Supplementary Figure 15: Manhattan plots of TWAS results of AD dementia by base training models. Significant genes are shown in orange and significant genes discussed in the main text are labeled and shown in red.**

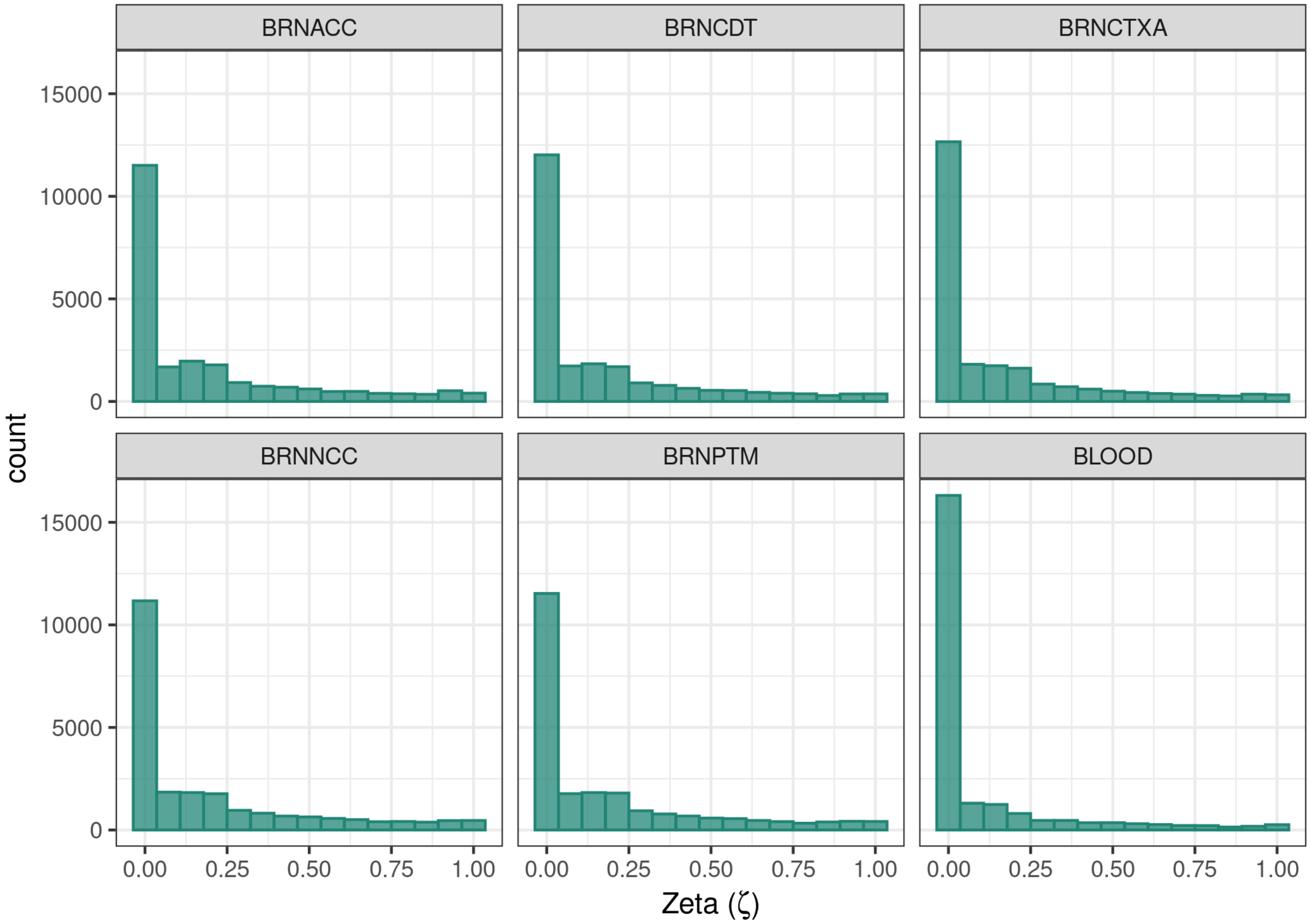

**Supplementary Figure 16: Plots of Zeta weights estimated by SR-TWAS for base training models used in PD TWAS.**

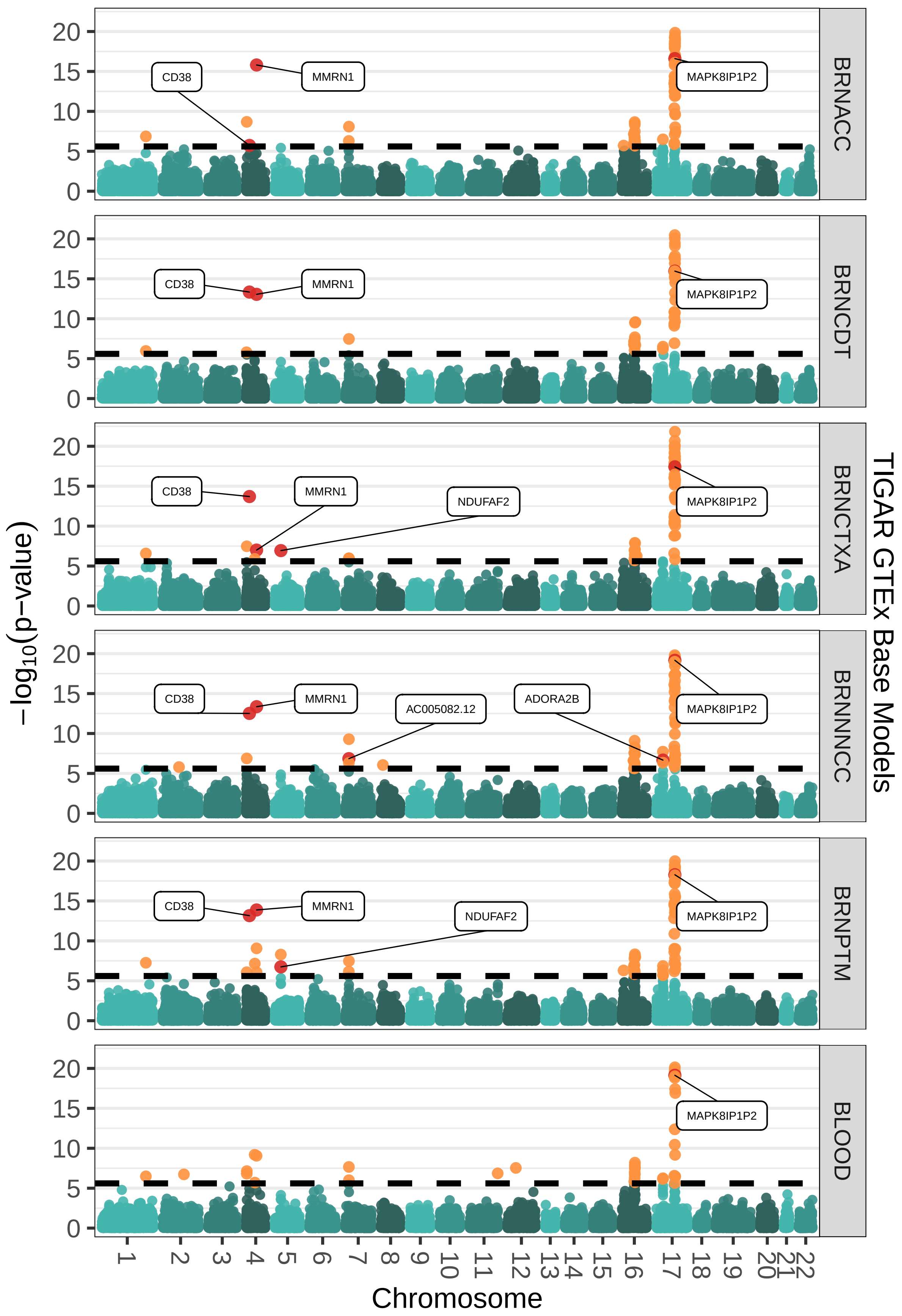

**Supplementary Figure 17: Manhattan plots of TWAS results of PD by base training models. Significant genes are shown in orange and significant genes that are discussed in the main text are labeled and shown in red.**
